# Plasma proteomics identifies early markers of endothelial and inflammatory activation associated with dengue disease severity in children

**DOI:** 10.64898/2026.03.15.26348146

**Authors:** Tatiana M. Shamorkina, Sofia Kalaidopoulou Nteak, Sokchea Lay, Ashwin Adrian Kallor, Sowath Ly, Veasna Duong, Albert J. R. Heck, Tineke Cantaert, Joost Snijder

**Author notes:** contributed equally, co-last, co-corresponding authors: Tineke Cantaert, Immunology Unit, Institut Pasteur du Cambodge, 1102 Phnom Penh, Cambodia., Joost Snijder, Bijvoet Centre for Biomolecular Research, Utrecht University, Utrecht, The Netherlands. These authors contributed equally to this work and share first authorship (Tatiana M. Shamorkina, Sofia Kalaidopoulou Nteak).

## Abstract

Dengue virus (DENV) is a major burden to global public health, affecting hundreds of millions annually. Children represent the major proportion of global dengue cases, ranging from asymptomatic or subclinical presentation to dengue fever (DF) and severe dengue hemorrhagic fever or shock syndrome (DHF/DSS). The factors that distinguish this range of disease severity are still poorly understood. To identify biomarkers of severity, we analyzed the plasma proteome of acute DENV infected children including both subclinical and hospitalized cases. Proteins associated with the acute-phase response, innate immune and lysosomal activation, and components of the coagulation cascade showed marked differences between hospitalized and subclinical cases during early infection. Longitudinal profiling demonstrated that endothelial dysfunction emerges early, with *PTX3* showing the strongest and most rapid upregulation in hospitalized patients, supporting its potential role as a marker of imminent vascular involvement. When comparing severe (DHF/DSS) and classical DF hospitalized cases, *CLEC11A* displayed the highest fold change at hospital admittance. We used machine-learning analysis to predict disease severity at the acute phase of infection, distinguishing subclinical from hospitalized cases and patients that develop classical dengue fever or severe disease based on the identified complement regulators and inflammatory markers. The panel of identified plasma proteins shed light on the mechanisms of dengue related disease progression and may provide a handle to predict disease severity based on blood markers present during the acute phase of infection.

## Introduction

Dengue is a systemic viral infection caused by the dengue virus (DENV) and transmitted primarily by *Aedes aegypti* mosquitos (1,2). DENV is a positive sense single-stranded RNA virus that belongs to the *orthoflavivirus* genus of the *Flaviviridae* family and further classified into four antigenically distinct serotypes (DENV1-4) (3,4). In 2019, dengue was listed among the World Health Organization’s top ten global health threats (2). Currently, half of the global population is at risk of dengue infection with an estimated 390 million cases annually in more than 120 countries, primarily in tropical and subtropical regions (5).

Infection with one of the four serotypes of DENV manifests as a clinical disease in 25-35% of cases ranging from classical dengue fever (DF) to the life-threatening dengue hemorrhagic fever (DHF) and dengue shock syndrome (DSS) (5,6). DHF/DSS, in which thrombocytopenia, hemorrhage, vascular leakage, and shock are the major clinical signs and possible cause of death, occurs predominantly after a secondary infection with a heterologous serotype (1,7). In endemic areas, children represent a major proportion of DHF/DSS cases (8,9) and their immune responses may differ from those observed in adults. Clinical progression typically follows four phases: acute (0-3 days after onset of symptoms), critical (3-7 days), early recovery (7-10 days) and recovery (∼90 days) (1,6). The clinical disease often begins with acute non-specific symptoms, including fever, headache, rash, and muscle or joint pain, with most patients recovering within 5-7 days (1). However, a sudden onset of plasma leakage 4-5 days after onset of symptoms (1,2,6) can lead to hypotension, cardiovascular collapse and as a result shock, and death (10). Currently, there is no antiviral therapy available for the disease, and the most effective treatment for dengue is hospitalization and careful administration of fluids (10,11).

Hallmarks of severe dengue include hyperinflammation and an aberrant immune response. Host-directed therapies could provide an interesting alternative for treatment of hospitalized dengue (12). However, the biological mechanisms mediating DENV control in the absence of disease manifestations remain poorly characterized, as the identification of asymptomatic or subclinical acute DENV-infected cases during routine surveillance is logistically challenging and resource intensive.

The viral secreted non-structural protein 1 (NS1) can contribute to the initiation of vascular leakage, a hallmark of dengue pathology, by directly disrupting the endothelial glycocalyx and compromising the tight junction integrity of the vasculature, thereby promoting vascular permeability (13). Furthermore, NS1 activates the immune system, fueling an exaggerated cytokine response that amplifies inflammation and endothelial damage (14). NS1 thereby creates a permissive environment for plasma leakage and intensifies the inflammatory response that precedes the onset of severe dengue. However, the molecular mechanisms underlying the transition from DF to severe DHF/DSS remain poorly understood, limiting the ability to predict progression before clinical deterioration becomes apparent. Indeed, during dengue outbreaks healthcare systems in low resource settings are restrained in capacity. In this context, early risk stratification of patients using predictive biomarkers is essential to prevent the development of hypovolemic shock and to reduce mortality associated with DHF/DSS (1,2,10).

Recent advances in plasma proteomics have enabled the identification of protein alterations associated with dengue pathogenesis (15). Multiple studies have attempted to define the molecular signatures distinguishing DF from DHF/DSS progression; however, these efforts have largely been limited to hospitalized adult cohorts, small sample sizes, single time-point sampling, or comparisons to healthy donors only, and in some cases relied on analytical approaches requiring highly controlled blood collection and processing conditions that are difficult to implement in dengue-endemic, resource-limited settings (16–23). Longitudinal proteomics analyses have nonetheless begun to highlight dynamic regulation of apolipoproteins, cytokines, endothelium regulation related proteins as well as immune markers across disease phases (16).

Here, we present a longitudinal plasma proteomics analysis of pediatric post-primary DENV-2 infections, spanning subclinical cases to hospitalized DF and DHF/DSS patients across all disease phases. By directly comparing subclinical and hospitalized dengue cases at matched time points, we identify protein-level differences that are already evident at the onset of symptoms. These early changes prominently involve pathways linked to endothelial activation, inflammation, and complement components, alongside broader virus-associated inflammatory signatures. Leveraging machine learning-based analysis, we built and tested a classifier for dengue disease severity to predict clinical outcomes from plasma biomarkers sampled early in infection. Together, this plasma proteomics dataset delineates early molecular features of dengue severity in children, laying a foundation for the development of early biomarkers applicable to pediatric dengue risk stratification.

## Results

### Clinical characteristics of the dengue pediatric cohort

The plasma proteomes of 92 participants, mostly children (Supplementary Table S1), with laboratory-confirmed post-primary DENV-2 infection and 10 healthy donors (age-matched children), all originating from Cambodia, were analyzed by reverse-phase liquid chromatography coupled to tandem mass spectrometry (LC-MS/MS). The cohort consisted of 23 subclinical cases (laboratory-confirmed dengue without meeting the WHO 1997 classification criteria for DF (24)), and 69 hospitalized patients (Figure 1A). The hospitalized dengue patients were classified according to the WHO 1997 classification criteria into DF, DHF and DSS (24). The plasma sampling was performed longitudinally across four disease stages: acute phase (AP; n=58 samples from hospitalized patients and n=15 from subclinical cases); critical phase (CP; n=60 hospitalized and n=8 subclinical); early recovery phase (ERP; n=60 hospitalized and n=19 subclinical) and recovery phase (RP; n=60 hospitalized and n=6 subclinical). In total, we analyzed 296 plasma samples of 92 dengue confirmed cases across all sequential phases of infection, in addition to 10 healthy donors. For the subclinical cases, samples were selected to correspond with the time points of the hospitalized patients based on onset of symptoms, ensuring that similar timepoints in the disease dynamics were compared between both groups. Both the dengue and healthy cohorts consisted of a balanced number of female and male individuals (dengue cohort: 48% females and 52% males, healthy donors: 50% females, 50% males). All laboratory-confirmed dengue cases were classified as post-primary infection with DENV-2 serotype using hemagglutinin inhibition test (HIA) on plasma obtained at day 0 and day 10.

**Figure 1.**
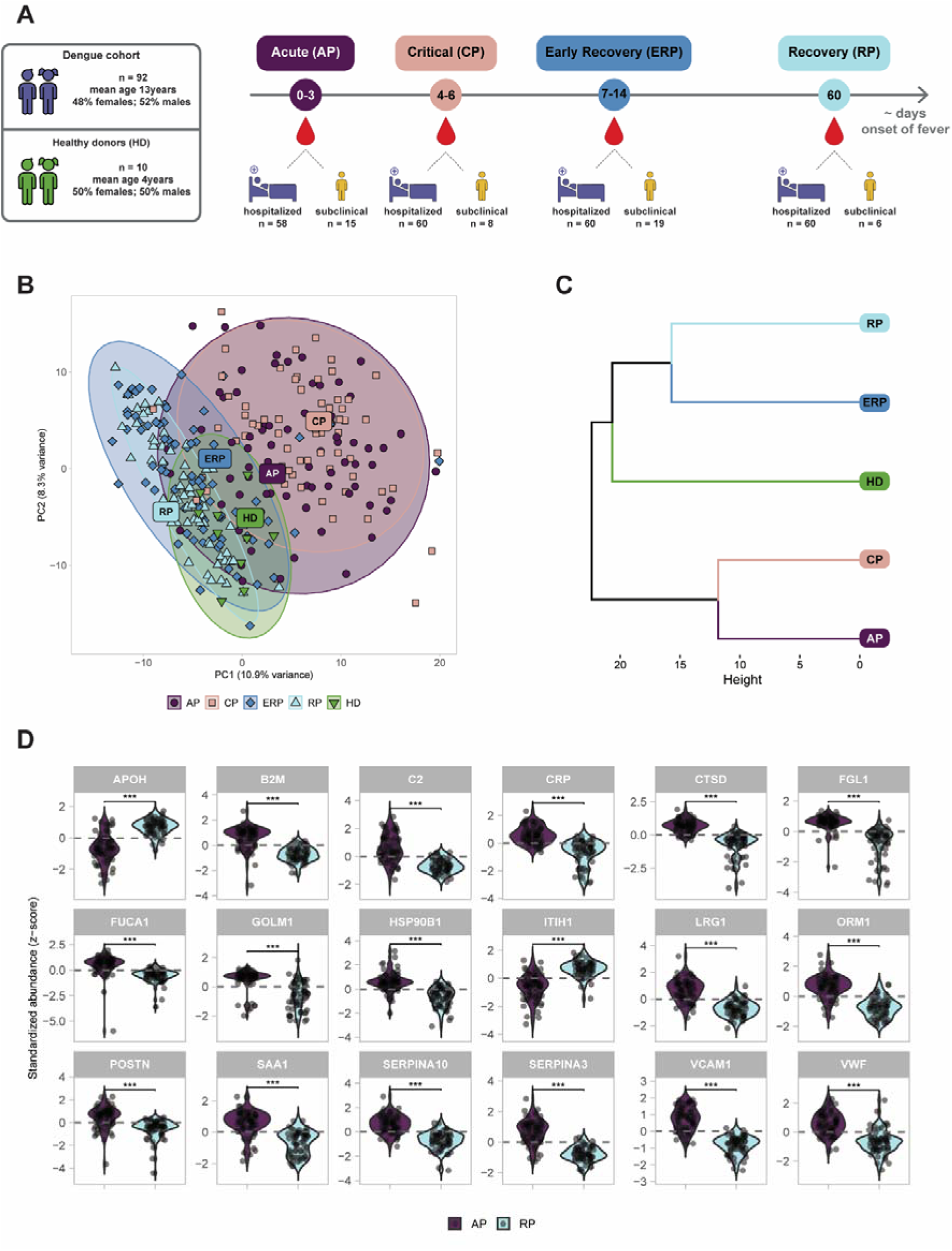
Plasma protein profile comparison of four clinical dengue disease phases observed in a pediatric cohort. A) Schematic representation of dengue pediatric cohort and plasma sample collection strategy from the acute phase (AP; n=73; in purple), the critical phase (CP; n=68; in pink), early recovery phase (ERP; n=79; blue), and the recovery phase (RP; n=66; light blue). Healthy donor group (HD; n=10) is represented in green. B) Principal component analysis (PCA) based on the plasma proteomics data of the 4 different phases of dengue infection (AP, CP, ERP and RP) and healthy donor group (n=10; HD). C) Clustering analysis of the different phases of dengue infection and healthy donor group. D) Violin plot shows the top 18 significantly regulated proteins contributing to the separation between AP (n=73; in purple) and RP (n=66; in blue) as revealed by using a random forest classifier. Statistical significance is shown above each protein and represented with stars (***: BH-adj p-value<0.001). Each dot within the violins represent the protein abundance value per individual.

### Plasma proteome profiles distinguish acute dengue infection from recovery stages in children

We could confidently quantify 500 proteins across all plasma samples (Supplementary Data S1). A panel of known red blood cell (RBC) contaminants, considered artefacts of sampling and sample preparation (25), as well as immunoglobulin variable domains, due to their sequence diversity and divergence from database sequences, were excluded from this dataset. Principal component analysis (PCA) based on all quantified proteins revealed a clear separation of the patients in the early phase of infection (AP and CP) from the recovery phases (ERP and RP) (Figure 1B). The patients in the acute and critical phases had closely related plasma proteome profiles, a pattern that was likewise observed between the early recovery and recovery phases. Moreover, the plasma proteome profiles of healthy donors showed minor interindividual variation, but generally overlapped with those of the recovering patients (Figure 1B), consistent with the expectation that two months post-infection marks the restoration of homeostasis after dengue infection (26). Similar patterns were observed in an unsupervised hierarchical clustering analysis (Figure 1C). Based on the branch height of the clustering dendrogram, the proteome profiles of AP/CP were closer to each other than those of ERP/RP. Based on our observations that healthy controls and recovery phase plasma proteomes were alike, we next focused exclusively on plasma proteome profiles from laboratory-confirmed dengue cases.

### Longitudinal plasma proteome profiling reveals inflammation-driven changes across dengue disease phases

Next, we compared the acute and recovery phase groups to track longitudinal changes in protein abundance over the course of dengue infection. Only proteins with Benjamini-Hochberg (BH) adjusted p-value <0.05 were considered significantly regulated (Supplementary Data S2). The top 18 differentially abundant proteins separating acute and recovery phase with adjusted p-value <0.001, summarized in Figure 1D, were identified by a mixed effects model and confirmed by a random forest classifier (Supplementary Figure S1). As expected, expression analysis demonstrated that the proteins differentiating acute dengue infection from the recovering patients reflected well-known signatures of inflammation (Figure 1D) and belonged to acute phase reactants (*CRP*, *SAA1*, *SERPINA3*, *ITIH1*, *ORM1, LRG1*) (27,28), markers of innate immune and lysosomal activation (*C2, CTSD, B2M, FGL1, FUCA1, HSP90B1*), signs of coagulation pathway activation (*SERPINA10, APOH*) (29), and proteins previously associated with viral infections (*GOLM1, APOH*) (30,31). The abundances of most significantly differentiated proteins increased during the acute phase whereas the abundances of *ITIH1* and *APOH* showed the opposite and were reduced (Supplementary Data S2). Furthermore, we observed an increase in the signatures of endothelial activation and vascular remodeling in the acute phase (*VCAM-1, VWF, POSTN, LGRG1*), which returned toward baseline levels during recovery (Supplementary Data S2). Taken together, these data indicate that longitudinal changes in the plasma proteome in dengue-infected cases can be mainly attributed to acute phase proteins, innate immune activation, coagulation and endothelial activation.

### Viral NS1 protein is detectable in plasma of laboratory-confirmed dengue cases

To determine whether dengue viral proteins were detectable in the plasma proteome and potentially could differentiate severity of the disease, we incorporated DENV-2 protein sequences into our proteomics database search. NS1 protein is secreted by DENV-infected cells and its detection is routinely used for the diagnosis of dengue infection during the acute phase (1). As expected, NS1 was absent in the plasma proteomes of healthy donors, but detected in plasma proteomes of 35 out of 92 laboratory-confirmed dengue cases (Supplementary Figure S2A). No other DENV-derived proteins beyond NS1 could be detected. NS1 was detected significantly more in the plasma proteomes of hospitalized severe patients (32/69, 46%) compared to subclinical cases (3/23, 13%, p=0.0057) (Supplementary Figure S2B), mostly during the acute phase, followed by the critical phase and in rare cases during the recovery period.

### Matched early stage plasma proteome differences distinguish hospitalized patients from subclinical dengue cases

We next compared plasma proteomic profiles between the subclinical dengue cases and hospitalized patients during the acute and the critical phases. Assessing these groups at matched early stages allows us to identify molecular changes in plasma of dengue infected individuals associated with a differential disease outcome. We found that 47 proteins were significantly altered in hospitalized patients across the acute and critical phases of the disease, particularly in the critical phase (Figure 2, Supplementary Data S3). Most proteins differentiating subclinical cases from hospitalized patients could be linked to inflammation and innate immune response, processes that typically accompany the onset of symptoms such as fever. A major subset of these pro-inflammatory proteins corresponded to acute phase reactants, including the canonical marker *CRP*, the similarly well-known *SAA2* and a range of other established acute phase proteins (*PTX3, ITIH2, ITIH3, ORM1, SERPINA3, SERPINF2, SERPING1, HRG*) (27,28,32). In our study, during the critical phase *PTX3* showed the strongest fold increase in hospitalized patients compared to subclinical cases, while *HRG* levels were reduced in the acute phase compared to subclinical cases. Moreover, acute phase reactants *ITIH2* and *ITIH3*, both inter-α-trypsin inhibitor family members, showed opposing regulation in hospitalized patients, with *ITIH2* exhibiting reduced and *ITIH3* elevated levels. We also observed *VCAM-1*, an established marker of endothelial activation (33), to be elevated in the hospitalized patients already during the acute phase. A substantial set of differentially regulated proteins in hospitalized dengue patients belonged to different arms of the host immune response (*CD14, ENPP2, LBP, COLEC11, LGAL3BP, WARS1, PVR, NCAM1, B2M, F5, C2, CFB, F13B, CFP, APOH*), many of which are released into the plasma upon infection (*CD14, NCAM1, ENPP2, GOLM-1, LGAL3BP, PVR, WARS1*) (31,34–39). Taken together, the proteins identified as differentially abundant during the acute and critical phases in hospitalized patients compared to subclinical cases reveal a rapid escalation of virus-driven inflammation and immune activation, aligning with the cytokine storm driven dengue immunopathology.

**Figure 2.**
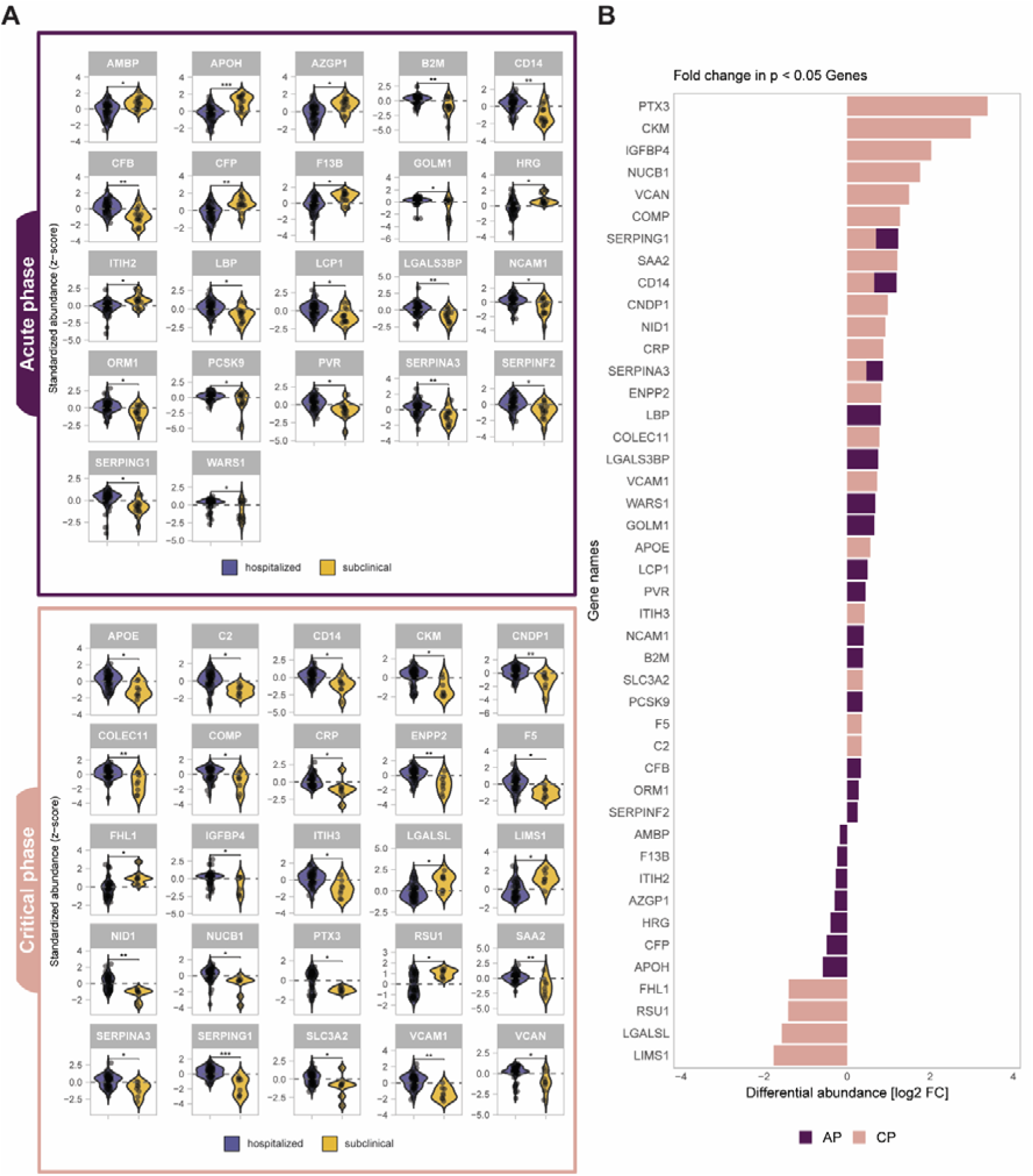
Plasma proteome profiles can distinguish hospitalized patients from subclinical cases in acute (AP) and critical phases (CP). A) Differentially abundant proteins between hospitalized patients (purple) and subclinical cases (yellow) in AP (top; purple panel) and CP (bottom; pink panel). Statistical significance is shown above each protein and represented with stars (*:BH-adj p-value<0.05; **: BH-adj p-value<0.01; ***: BH-adj p-value<0.001). Each dot within the violins represent protein abundance value per individual. B) Fold change (FC) in plasma abundance of differentially regulated proteins from panel A. Positive FC shows that the proteins are higher abundant in the hospitalized patients . The bars for proteins that are significantly enriched in AP or CP are colored in purple and pink, respectively. Stacked bars show that these proteins were significantly changed in plasma abundance in both phases.

### Dengue disease symptoms onset signals early endothelial stress, led by early acute-phase reactant PTX3 increase and pro-inflammatory changes in hospitalized patients compared to subclinical cases

Dengue is a dynamic disease, where both biological and clinical changes rapidly occur (40). Hence, we evaluated the temporal evolution of the differential proteins in hospitalized patients versus subclinical cases across all four sampling phases of the disease. Based on the functional annotation using literature and Uniprot database, the differentially abundant proteins identified in Figure 2 naturally fell into two prominent biologically relevant categories: endothelial dysregulation and viral-induced inflammation. For these proteins, we plotted the mean protein abundance (LFQ) across all timepoints in hospitalized and subclinical cases (Figure 3) to characterize how protein abundances evolved across dengue disease phases. Among these molecules, *CD14* (monocyte marker) and *LBP* (LPS binding protein), previously described as elevated in severe dengue patients (41), showed distinct temporal patterns in our study. *CD14* was already elevated in the acute phase, reflecting early monocyte activation, whereas *LBP* reached its highest levels during the critical phase. In addition, several complement-related proteins displayed phase-specific patterns (Figure 3), with elevated levels of *C2* and the complement activator *COLEC11* during the acute phase, followed by increased *CFB* during the critical phase in hospitalized patients.

**Figure 3.**
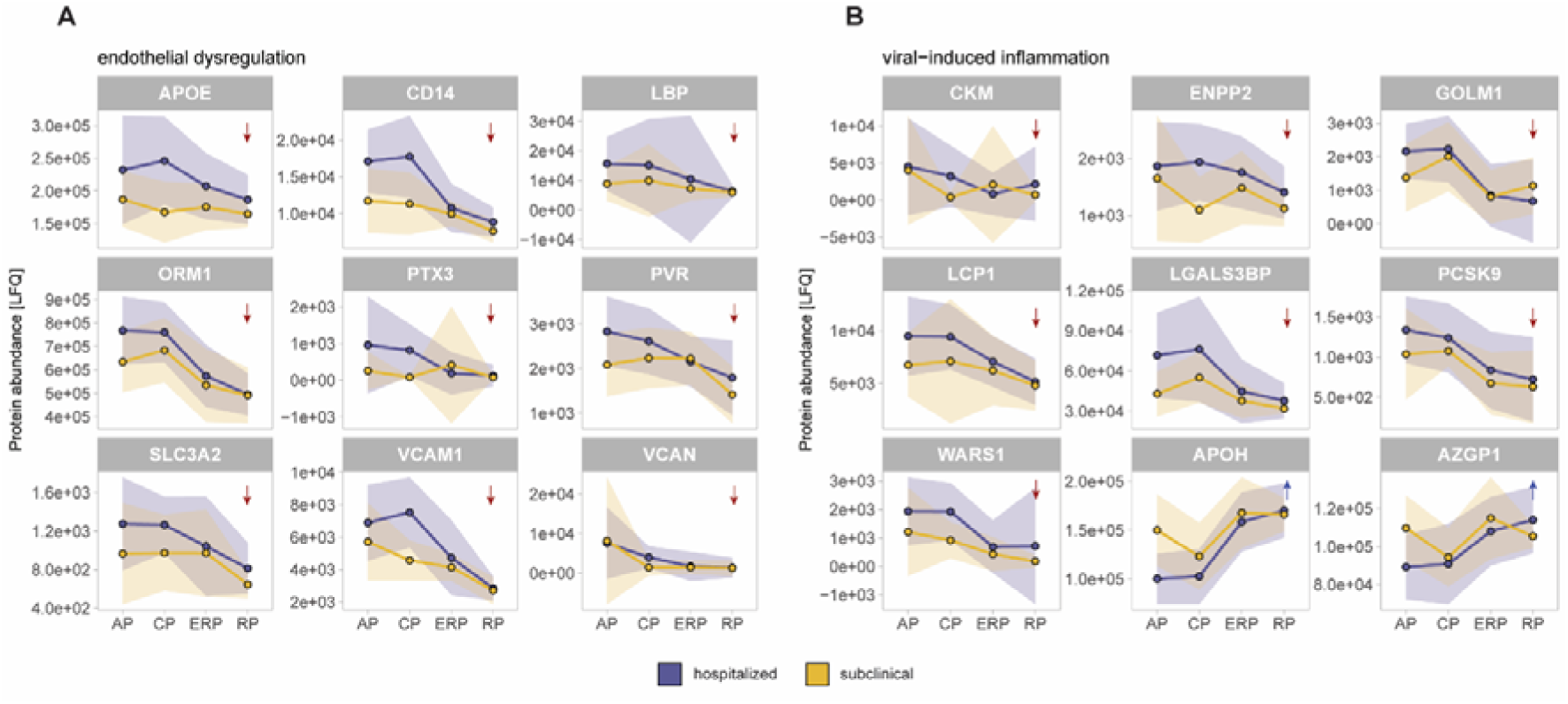
Distinct longitudinal trends in hospitalized patients and subclinical cases for proteins involved in A) endothelial dysregulation or B) viral-induced inflammation. The proteins in both panels were chosen based on the functional annotation in literature and Uniprot from the differentially abundant proteins shown in Figure 2. The solid lines show the average protein abundance (LFQ) of each protein in each phase, and the faded purple or yellow areas indicate the standard deviation of each phase in the hospitalized patients or subclinical cases, respectively. The red or blue arrows on the top-right corner of each plot reveal increasing or decreasing trends. The trends are shown through all dengue disease phases.

One of the major hallmarks of clinical dengue disease pathogenesis, particularly in severe cases, is hemorrhage, driven by the disrupted glycocalyx and plasma leakage across the compromised endothelial barrier (1,2,13). We observed that a substantial number of proteins elevated in hospitalized patients during the acute phase were associated with endothelial integrity and capillary barrier function (Figure 3A). These proteins (*APOE, VCAM-1, CD14, LBP, VCAN, PTX3, PVR, ORM1 and SLC3A2*) consistently increased during both the acute and critical phases and returned to baseline levels during the recovery. A few proteins (*APOE*, *LBP, SLC3A2*) showed a broader variation even in the recovery stage. Although their changes were most pronounced in hospitalized patients, these proteins also exhibited measurable shifts in subclinical cases, though to a lesser extent, across the acute and critical phases. These findings indicate that endothelial barrier dysregulation can be observed in both subclinical and hospitalized dengue patients and is already prominent at the onset of clinical dengue symptoms, with the pentraxin *PTX3* emerging as the earliest and strongest responder.

Another major hallmark of dengue pathogenesis is virus-driven inflammation. Accordingly, our data revealed a panel of differentially abundant proteins linked to virus-induced inflammation (Figure 3B) documented in previous viral infection studies (28–31,35,36,42–47). Most of these proteins (*CKM*, *ENPP2*, *LCP1*, *LGALS3BP*, *GOLM1, PCSK9* and *WARS1*) were increased in hospitalized patients compared to the subclinical cases during the acute and critical phases, whereas the levels of *APOH* and *AZGP1* were consistently reduced in the hospitalized patients during the same phases of the infection. All proteins except *WARS1* returned to baseline levels in the recovery phases, while *WARS1* remained high even 60 days after onset of fever, as reflected by the high standard deviation observed among hospitalized patients in Figure 3B. These data indicate that DENV infection extensively alters host proteins required for mounting and regulating antiviral immunity, with these disruptions being far more pronounced in the acute phase of hospitalized patients compared to subclinical cases.

### Plasma proteome signatures differentiate DF from DHF/DSS pediatric patients at hospital admittance

To identify prognostic markers of severity at hospital admittance, we investigated differences in the plasma proteomes between hospitalized DF and DHF/DSS patients during the acute phase of dengue, searching for early differentiating markers of disease severity. We performed a two sample *t*-test using a gene-wise linear regression model with clinical classification (DF vs DHF/DSS) as the independent variable. We identified a panel of 18 proteins with significantly different abundances in the plasma of the DHF/DSS patient group in comparison to the DF patients (Figure 4 and Supplementary Data S4). Some proteins (*COLEC11*, *CFP*, *APOE*, and *LCP1*), observed at elevated values in the DHF/DSS group were also seen as markers of disease when compared to the subclinical cases (Figure 2).

**Figure 4:**
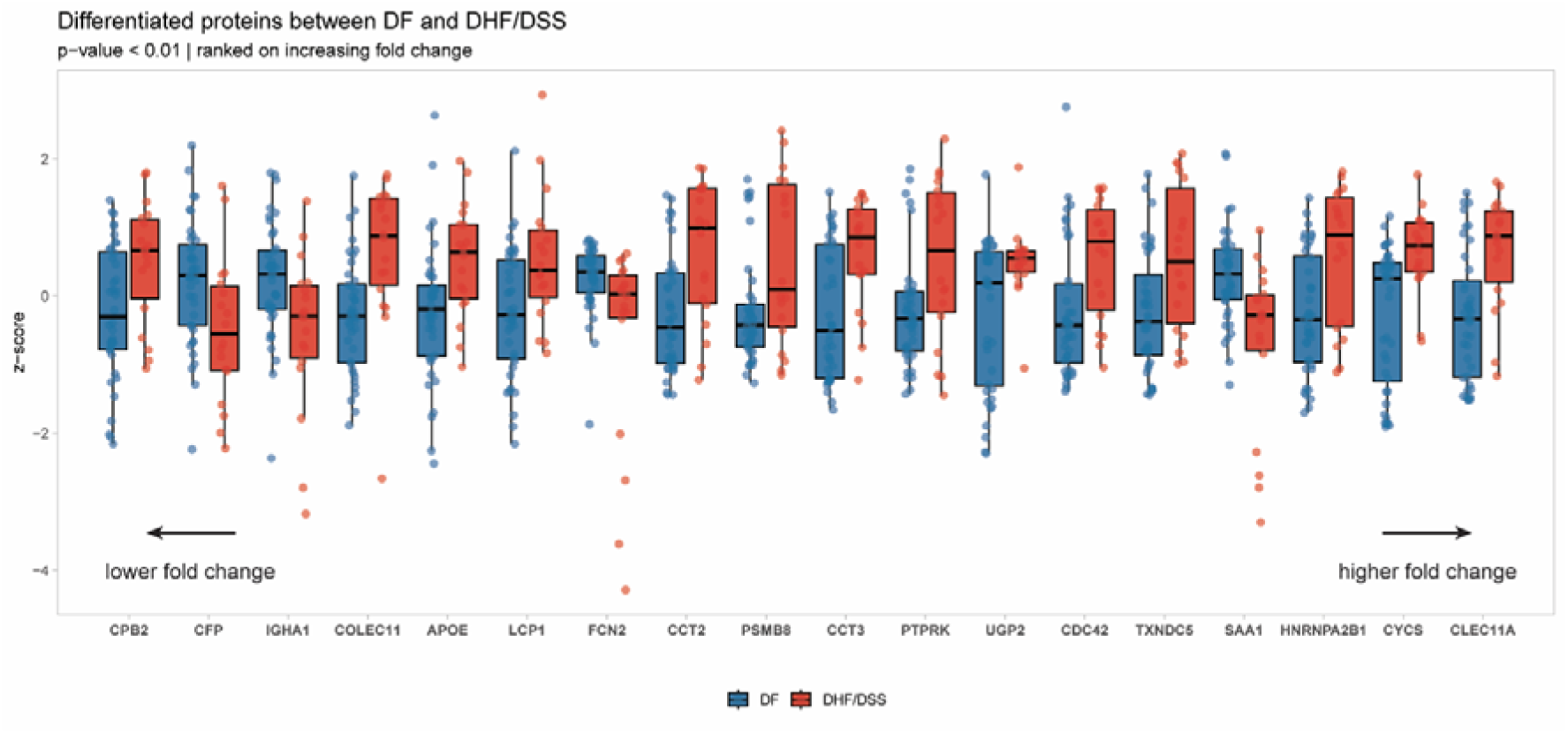
Plasma proteins with significantly different abundance between DF (blue) and DHF/DSS (red) patients in the acute phase (AP). Proteins are ordered from low to high fold change, meaning that CLEC11A displayed the highest difference in abundance between the two conditions in the acute phase. The values are represented in z-score calculated from the protein abundance values to the median. All shown proteins passed a two sample t-test using a gene-wise linear regression model with p-value<0.01.

Among the proteins differentiating DHF/DSS from DF patients, *CLEC11A* exhibited the largest fold change. In addition, differentially abundant *FCN2*, *CFP* and *COLEC11* proteins represent two different segments of complement pathway (48). *CFP* is increased in DF patients and stabilizes the alternative pathway of complement activation while *FCN2* and *COLEC11* are increased in DHF/DSS patients and are involved in the lectin pathway. These data indicate a dysregulation of the complement pathway activation during severe disease. Approximately half of the differentially abundant proteins (*CCT2*, *CCT3*, *CDC42*, *PSMB9*, *HNRNPA2B1, TXNDC5* and *CYCS*) were cytosolic host factors, previously implicated in the cellular stress response and are known to act as host-dependency factors for multiple viruses (49–55). Their release into plasma of severe dengue patients likely reflects infection-induced cellular damage and overall viral burden. Finally, total IgA1 levels were significantly lower in DHF/DSS patients compared to DF cases, raising the possibility that an insufficient IgA response may contribute to progression to severe dengue.

### Changes in antibody subclasses during dengue infection

Observing that DF and DHF/DSS patients differed in IgA levels, we extended our analysis to examine antibody subclasses more broadly and compared their abundances across hospitalized patients, subclinical cases, and healthy controls throughout all phases of dengue disease progression. In our study, the total abundances of antibody isotypes (IgM, IgG, and IgA) remained largely unchanged across dengue disease states and recovery phases in all infected individuals (Supplementary Figure S3). However, IgA levels were significantly increased in infected individuals compared to healthy donors across all phases of infection (Supplementary Figures S4-S7). In addition, we observed significant changes in the abundances of IgG1, IgG2 and IgG4, along with a notable upward trend in IgG3 levels in the infected individuals compared to healthy donors during the acute phase (Supplementary Figure S4). IgM levels stayed stable throughout the disease progression phases and in comparison to the healthy controls, possibly due to the post-primary infection status (Supplementary Figures S4-S7).

### Classification of patients using machine learning models provide a panel of putative biomarkers of dengue severity in the acute phase of the disease

Building on the plasma proteomics analysis across disease states above, we next developed machine learning classifiers to assess whether dengue disease state and the patient progression could be predicted at the acute phase of dengue disease. The classifiers were built using the panel of significant differentially abundant proteins identified by the mixed-effects model in the preceding analyses (Figures 2-3). Area under the receiver operating characteristic curve (AUROC) analysis using leave-one-pair-out cross-validation (LOPO-CV) revealed strong classifier performance across all three classification tasks: acute versus recovery phase, hospitalized versus subclinical, and dengue fever versus dengue hemorrhagic fever (Figure 5). The classifier achieved the highest performance in distinguishing acute from recovery phases (AUROC = 0.992, N = 79) (Figure 5A, left), followed by hospitalized versus subclinical patients (AUROC = 0.887, N = 73) (Figure 5A, center), and dengue fever versus dengue hemorrhagic fever (AUROC = 0.762, N = 58) (Figure 5A, right).

**Figure 5:**
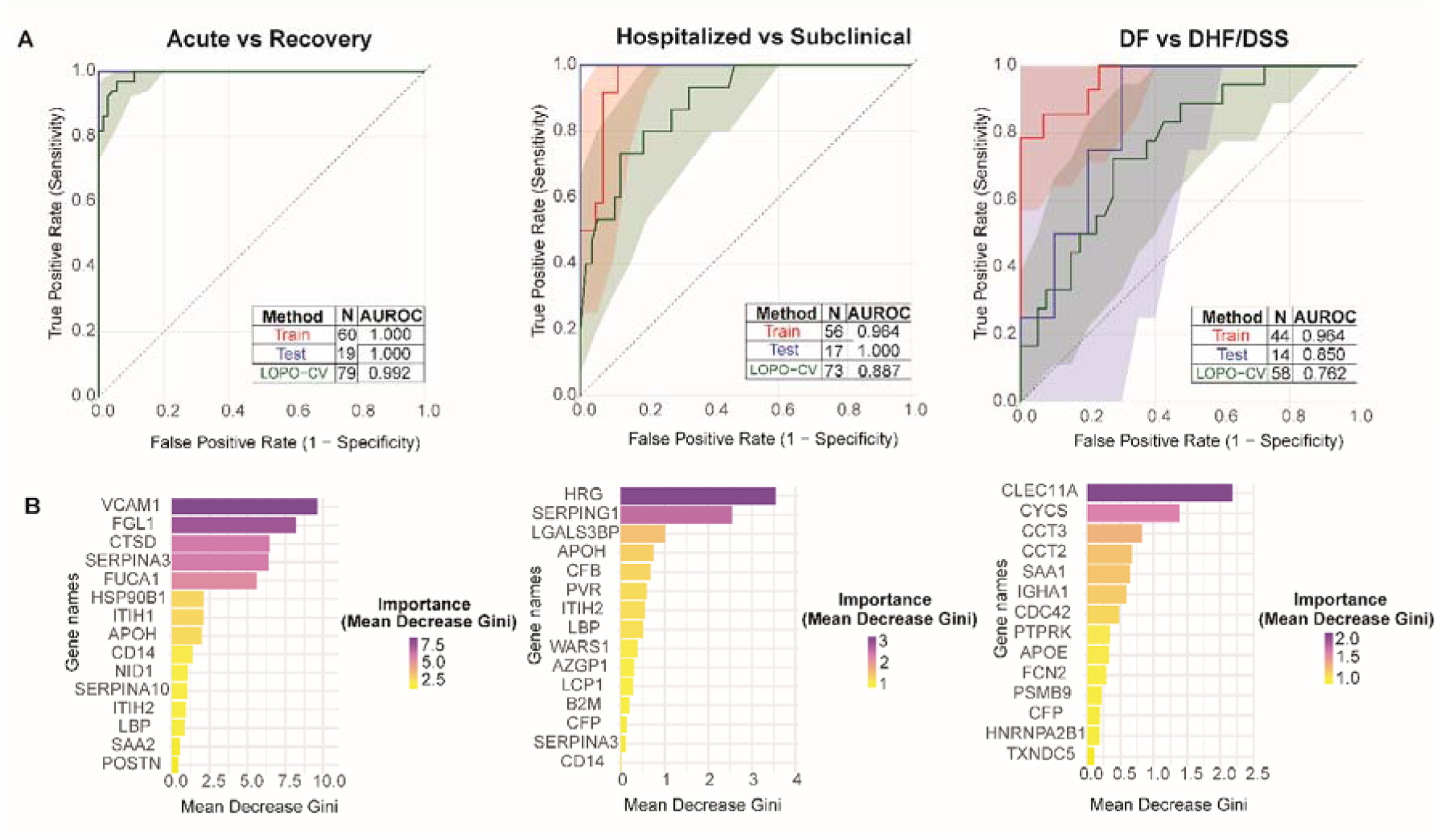
Random forest classifier results for the discrimination of the patients in different classes in the acute and recovery phases of dengue disease. A) The performance metrics of random forest classifiers distinguishing acute and recovery phases (left), hospitalized and subclinical (center) and dengue fever and dengue hemorrhagic fever (right). The random forest models achieve strong discrimination across comparisons based on their Leave-One-Patient-Out Cross Validation (LOPO-CV) Area Under the Receiver Operating Characteristic Curve (AUROC) values. B) The top 15 features ranked by mean decrease in Gini index, a measure of each variable’s contribution to classification purity. Among the genes determined to be the most discriminative are VCAM1 and FGL1 for the acute vs recovery phase classification, HRG and SERPING1 for the hospitalized vs subclinical classification and CLEC11A and CYCS for the dengue fever vs dengue hemorrhagic fever classification. (Mean decrease gini measures how much a gene influences a split between the two classes; the higher the value, the more the gene influences the split between the two classes).

Besides assessing model performance, the genes that were important in distinguishing the various classes were identified through feature importance analysis. Feature importance analysis revealed distinct molecular signatures for each classification task (Fig 5B). In distinguishing acute from recovery phases, VCAM1 showed the highest importance (mean decrease in Gini ∼10.0), followed by FGL1, CTSD, SERPINA3, and FUCA1 (Figure 5B, left). For the hospitalized versus subclinical classification, HRG emerged as the strongest discriminative feature (mean decrease in Gini ∼4.0), with other top contributors including complement pathway proteins SERPING1, LGALS3BP, APOH, CFB, and PVR. The prominence of SERPING1, a major inhibitory regulator of the complement system, indicates early complement activation in clinically apparent disease (Figure 5B, center). The separation of DF from DHF/DSS patients in the acute phase was dominated by CLEC11A (mean decrease in Gini ∼2.5), followed by CYCS, CCT3, CCT2, SAA1, IGHA1, and CDC42. This signature includes an early acute phase reactant (SAA1), cytosolic proteins (CYCS, the CCT2/CCT3 TRiC complex, and CDC42), as well as IGHA1. A complementary logistic regression approach with elastic net regularization yielded largely concordant results, albeit with different rankings of the discriminating features (Supplementary Figure S1). Taken together, these results suggest that plasma proteome signatures measured during acute dengue infection can accurately predict disease trajectory and severity, with distinct molecular profiles distinguishing disease phases, clinical presentation, and progression to severe manifestations.

## Discussion

Our study of pediatric post-primary DENV-2 infections establishes endothelial dysfunction and dysregulated inflammation as the earliest and most pervasive drivers of dengue disease. Hospitalized patients showed coordinated increases in inflammatory and endothelial-associated plasma proteins during the acute phase, which persisted into the critical phase and largely normalized during recovery. These longitudinal changes were dominated by acute phase reactants (*CRP*, *SAA1*, *SERPINA3, ITIH1, ORM1, LRG1*), markers of innate immune and lysosomal activation (*C2, CTSD, B2M, FGL1, FUCA1*, *HSP90B1*), proteins linked to coagulation and complement regulation (*SERPINA10, APOH*), and factors previously associated with viral infection responses (*GOLM1, APOH, PCSK9*). In contrast to the general pattern of upregulation, *ITIH1* and *APOH* were reduced early in infection and normalized during recovery. Notably, *APOH* is known to bind viral particles and promote their clearance via phagocytosis (56,57), which is in line with *APOH* transient depletion during acute disease seen in this study. The subsequent restoration of *APOH* abundance during recovery is consistent with resolution of viral burden and inflammatory drivers.

In parallel, proteins indicative of endothelial activation and vascular remodeling, including *VCAM1, VWF, POSTN* but also acute phase proteins like *ORM1*, and *LRG1*, were elevated during acute disease and declined during recovery. Endothelial activation represents a pro-inflammatory state in which endothelial cells upregulate adhesion molecules, alter barrier integrity, and release mediators that promote leukocyte recruitment and glycocalyx disruption (58). The coordinated regulation of the identified markers underscores vascular injury as a central determinant of DENV progression.

Endothelial dysregulation also emerged as a dominant early feature distinguishing hospitalized patients from subclinical cases. Proteins associated with endothelial activation, barrier remodeling, and glycocalyx integrity, including previously mentioned *VCAM-1* and *ORM1*, but also *VCAN*, *CD14*, *LBP, ITIH2/ITIH3, PTX3, SLC3A2, PVR/CD155* and *APOE* were elevated during the acute and critical phases in hospitalized patients in comparison to subclinical cases. *VCAM-1*, which promotes leukocyte adhesion and monocyte-endothelium bidirectional crosstalk (59), was increased in parallel with soluble *CD14*, a marker of aberrant monocyte activation previously linked to plasma leakage severity and overall dengue severity (38,41,60). These results further highlight the early immune activation at the vascular interface. Elevated *VCAN*, a cytokine-induced extracellular matrix proteoglycan synthesized by endothelial cells (61), in hospitalized patients reflects a cytokine-driven extracellular matrix remodeling that facilitates leukocyte infiltration (61) while *PTX3* exhibited the highest fold change and has been associated with endothelial glycocalyx disruption through its correlation with circulating SDC1 (19,62). Additional endothelial surface proteins in our study, *SLC3A2* and *PVR/CD155*, point to altered immune regulation at the vascular surface in hospitalized dengue (45,63). Interestingly, proteins with endothelium protective functions, including *ORM1* and *APOE*, were also increased, suggesting activation of compensatory mechanisms. *ORM1* is an acute phase reactant that is known to limit vascular leakage by modulating endothelial permeability (64), whereas *APOE* possesses anti-inflammatory and endothelium repair properties. However, *APOE* has been also shown to bind circulating NS1 in plasma and accumulate as an *APOE-*NS1 complex during the critical phase (42), potentially attenuating its protective function. Together, these patterns indicate that endothelial barrier dysregulation is established at the earliest symptomatic stages of dengue, driven by an imbalance between inflammatory injury and host compensatory responses.

These endothelial dysregulation signatures are accompanied by broad activation of innate immune and complement pathways. Hospitalized patients displayed coordinated increases in acute-phase proteins, complement components, and immune-regulatory molecules in the acute disease in comparison to subclinical cases. The regulation of complement proteins, characterized by higher *C2* and *COLEC11* in the hospitalized patients during the acute phase and higher *CFB* during the critical phase in the same patients, suggests a dynamic shift from classical pathway engagement that is present in the subclinical cases toward alternative pathway amplification in the hospitalized patients (48,65–67). Such complement dysregulation has been implicated previously in severe dengue disease and is consistent with immune driven vascular pathology (48,67).

Beyond endothelial and complement pathways, we observed dysregulation of several host proteins associated with virus-induced inflammation and cellular stress, including *ENPP2, AZGP1, WARS1, LCP1, APOH, LGALS3BP*, *PCSK9* and *GOLM1*. Many of these proteins have been reported as markers of disease severity or hyperinflammation in other viral infections, pointing to conserved host responses across distinct viral pathogens (28–31,35,36,42–47,68). For example, higher concentrations of *APOH*, *AZGP1* and *LCP1* have been correlated to poor prognosis after viral infections, including infections with Influenza and SARS-CoV-2 viruses (30,46). *WARS1* and *ENPP2* have earlier been associated with elevated levels of pro-inflammatory cytokines such as IL-6 while *ENPP2* also correlated with elevated levels of secreted *CD14* described above (36,37,47). *LCP1*, a known critical regulator of immune cell function, has similarly been found increased in severe COVID-19 patients (46,69). DENV was previously shown to induce *PCSK9* levels in plasma of patients with high viremia (68). On the other hand, *GOLM1*, a component of the host cell Golgi/secretory machinery (31), has been shown to promote hepatitis C virus secretion, while DENV is known to induce a Golgi stress response to enhance virion processing and secretion (70).

Notably, several proteins in our cohort distinguishing hospitalized patients from subclinical cases have previously been associated with mortality or adverse outcomes in other severe inflammatory conditions. Two acute-phase proteins, *PTX3* and *HRG*, showed opposing regulation in hospitalized patients, with *PTX3* markedly elevated and HRG reduced. Both proteins have been linked to mortality in prior studies, including COVID-19, albeit with contrasting functional implications (27,71–73). In addition, *WARS1*, which was increased in hospitalized patients, was previously also reported as an early mortality associated marker in sepsis patients and correlated with severe systemic inflammations and organ damage (37). The persistence of *WARS1* dysregulation into recovery further suggests that hospitalized patients experience prolonged post-acute inflammatory remodeling, even after clinical resolution. Furthermore, increased *CKM* (creatine kinase) abundance in hospitalized patients indicates muscle tissue damage and increased vascular permeability, in line with prior clinical dengue studies that reported elevated plasma *CKM* levels (74,75).

The patterns of endothelial activation, vascular dysfunction and inflammation are further supported by our detection of circulating viral NS1 protein in plasma, predominantly in hospitalized DHF/DSS patients and most frequently during the acute phase, coinciding with peak endothelial stress and inflammatory markers. Although NS1 was detectable only in a subset of laboratory-confirmed cases and at low abundance, its identification demonstrates that viral proteins can be captured within untargeted plasma proteomic datasets. The NS1 derived tryptic peptides observed here represent measurable viral signals in circulation and suggest that targeted mass spectrometry approaches could enable more sensitive quantitative NS1 detection, as previously demonstrated for SARS-CoV-2 spike peptides measured in nasal swabs (76,77), complementing existing immunoassay-based diagnostics.

Early risk stratification of dengue patients at hospital admission remains a major unmet need. In our study, differences between DF and DHF/DSS patients at admission highlighted pathways associated with disease severity, including markers of activation of the alternative and lectin complement pathways (*COLEC11, CFP, FCN2*), endothelial stress (*APOE*), and immune regulation (*LCP1*), which were enriched in DHF/DSS patients. *CLEC11A* showed the strongest stratification of severe cases and has been reported as a circulating marker of hyperinflammatory states in other viral diseases (78). *CLEC11A* (Osteolectin) promotes osteogenesis under inflammatory conditions (79), but its role in dengue remains to be elucidated. Approximately half of the discriminating proteins were cytosolic host factors implicated in cellular stress responses and viral dependency, likely reflecting increased cellular damage. In addition, lower IgA1 levels in DHF/DSS compared to DF patients suggest that an insufficient IgA response may accompany progression toward severe disease. The role of IgA during DENV infection remains unclear, as DENV-specific IgA can neutralize DENV without inducing ADE, while anti-NS1 IgA can induce neutrophil activation during secondary flavivirus infection (80,81). Elevated IgA and broader antibody subclass abundances across all dengue cases relative to healthy donors further indicate a rapid humoral response following post-primary infection.

Beyond defining pathophysiological trends of dengue disease, we developed machine-learning based classifiers to address the differences driving the acute and recovery phases, subclinical cases from hospitalized patients as well as the independent stratification of the DF and DHF/DSS patients. We identified a robust and convergent set of plasma proteins, including mortality markers *HRG* and *WARS1* (28,37,72), as well as complement pathway proteins *SERPING1*, *CFB* and *APOH* that distinguished subclinical from hospitalized patients in the acute phase, while inflammatory markers such as *VCAM1* and *FGL1* most effectively distinguished acute from recovery phase patients. Notably, discrimination between DF and DHF/DSS was driven by *CLEC11A* alongside cytosolic stress-associated proteins, including *CYCS* and TRiC complex components (*CCT2/CCT3*), *IgA1* and an early acute-phase reactant *SAA1*, suggesting that severe disease is characterized by heightened cellular stress, tissue injury and early immune response already at hospital admission.

In summary, this longitudinal plasma proteomics study demonstrates that the difference between subclinical to clinical dengue in children is marked by early and coordinated activation of endothelial stress, complement pathways, and innate immune responses. By capturing these processes across disease phases and clinical presentation, our data refine current models of dengue immunopathogenesis and highlight candidate biomarkers that may support early risk stratification and improved clinical management in pediatric dengue.

## Conclusions

Our findings expand the molecular understanding of dengue severity in children and demonstrate the power of time-resolved clinical proteomics to capture both viral and host determinants of disease progression. These protein signatures highlight potential druggable targets and offer promising opportunities for the development of early risk-stratification tools.

## Limitations of this study

Although sampling spanned all clinical phases of dengue disease, complete longitudinal profiling was available for only a subset of participants (n = 11), limiting our ability to resolve person specific temporal trajectories with high confidence. Second, identification of acute infected subclinical cases is logistically challenging and resource consuming, and while this work provides the first plasma proteomic characterization of subclinical dengue infection cases, the number of subclinical samples was considerably smaller (n = 15 for the acute phase) than the hospitalized cohort (n = 58 for the acute phase). This may introduce imbalance and reduce statistical power. Finally, this study focuses on pediatric patients from a single geographic region infected with a single DENV serotype, which may limit generalizability to adult populations or other settings. However, children represent a major and vulnerable group affected by severe dengue, underscoring the relevance of these findings. To generalize our results, the panel of identified markers should be validated in cohorts of patients with other genetic background and different DENV infecting serotypes.

## Materials and Methods

### Ethics statement

The study was performed in accordance with the principles of the Declaration of Helsinki. Ethical approval for the clinical study was obtained from the National Ethics Committee for Health Research of Cambodia (NECHR nr 2020-214). Written informed consent was obtained from all participants or from the legal guardians of participants under 16 years of age prior to inclusion in the study.

### Patient recruitment

DENV cases were identified in the DENTHOM project from 2022 to 2024 among hospitalized patients presenting with acute dengue-like illness in Kampong Thom and Siem Reap provinces. Patients were recruited at Kampong Thom Provincial Hospital, Baray and Stuong district hospitals in Kampong Thom province and Jayavarman VII hospital in Siem Reap province. Blood samples were collected upon hospital admission (day 0, D0) and tested for DENV by quantitative reverse transcription polymerase chain reaction (RT-qPCR) (82), day 3 (D3), day 10 (D10) and day 60 (D60) after admission. Hospitalized patients were classified according to the WHO 1997 classification scheme into dengue fever (DF), dengue haemorrhagic fever (DHF), or dengue shock syndrome (DSS) (83). To identify individuals with subclinical infections, a household investigation approach was used around RT-qPCR-confirmed index cases in Kampong Thom province. In laboratory-confirmed cases, clinical signs and symptoms were recorded retrospectively for 5 days and prospectively for 10 days, with follow-up visits on days 3, 5, and 10 (D3, D5, and D10) post-enrolment. Subclinical infection was defined as no or mild (<38 °C) fever for a maximum of 2 consecutive days and a maximum of 1 of the following symptoms for a maximum of 2 consecutive days: nausea or vomiting, rash, aches and pains. Individuals requiring medical care were excluded from subclinical cases. For all RT-qPCR-confirmed DENV cases, additional blood samples were collected on days 3, 10, and 60 after inclusion. Clinical and biological data were documented in case report forms. Samples from both hospitalized and subclinical cases were categorized by infection stage according to WHO2009 guidelines (84) in order to ensure similar timepoints in the disease dynamics was compared between both groups: acute phase (0-3 days after onset of fever), critical phase (4-6 days after onset of fever), early recovery (7-14 days after onset of fever) and recovery (2 months post onset of fever). In the subclinical group who had no or mild symptoms, D0 and D3 samples were categorized in the acute and critical phases, respectively. Healthy donors were identified in household samples and defined as DENV RT-qPCR negative

### Laboratory diagnosis

Plasma samples from patients were tested for the presence and serotype of DENV by RT-qPCR at the Virology Unit of the *Institut Pasteur du Cambodge* (IPC) as described (82). The hemagglutination inhibition assay (HIA) was used to distinguish between primary and post-primary infection (84) and only post-primary infections with DENV-2 were included in the subsequent analysis

### Plasma sample preparation for proteomics

The plasma proteomics workflow was adapted from a previous study (85). Briefly, 10 µL of plasma samples were diluted 2-fold with 100 mM Tris buffer, pH 8.5. 4 µL of the diluted samples were mixed with 78 µL of lysis buffer (4% sodium deoxycholate (SDC); 160 mM chloroacetamide (CAA); 100 mM Tris, pH 8.5; 40 mM tris(2-carboxyethyl)phosphine (TCEP)). The mixture was incubated at 95°C for 10 min for denaturation and reduction/alkylation. Next, the samples were further diluted with 100 µL of 100 mM Tris buffer, pH 8.5 and 10 µL of Trypsin (Sigma-Aldrich) and LysC (Wako) at 0.2 µg/µL stock concentration were added to each sample. After the overnight digestion at 37 °C, the enzymatic activity was quenched with the addition of 200 µL of 2% trifluoroacetic acid (TFA). The resulting peptides was further diluted 36-fold in 1% TFA.

### LC-MS/MS DIA for proteomics

20 µL containing approximately 200 ng of peptides, were loaded onto Evosep® Pure tips according to Evosep manufacturer protocol and measured on an Evosep One LC system (Evosep, Odense, Denmark) coupled to a timsTOF HT mass spectrometer (Bruker Daltonics, Bremen, Germany). The data was acquired in dia-PASEF-MS mode. The peptides were eluted from the Evosep® Pure tips followed by the peptide separation on an EV-1109 analytical column (ReproSil Saphir C18, 1.5 μm beads by Dr Maisch; 8 cm length × 150 μm i.d.; Evosep) using the 60 SPD method. Elution gradient was realized using mobile phases A (0.1% formic acid in Milli-Q water) and B (0.1% formic acid in 100% acetonitrile). Peptides were ionized with captive sprayer (ZDV, 20 µm ID, 1865691, Bruker Daltonics) that was operated at 1,600 V, with drying gas flowing at 3 l/minute and drying at 180°C. The dia-PASEF-MS mode and ion mobility (IM) windows were configured as previously described by Michalik et al (85). Peptides were isolated within the m/z range of 300-1200, with 12 dia-PASEF scans and 8.3% duty cycle. The IM range was set to 1.6 and 0.6 V cm2; the accumulation and ramp times were set as 100 ms. Collision energy ramping was set to default (20-59 eV within the mobility range of 0.6-1.6 Vs/cm2). Acquisition was conducted with a ramp and accumulation time of 100 ms, achieving a 100% duty cycle.

### Raw data processing and statistical analysis

Raw files were searched using DIA-NN (version 1.9) in library-free mode (86). Trypsin was selected as the protease and two missed cleavages were tolerated. Cysteine carbamidomethylation and methionine oxidation were allowed. The mass accuracy was set to 20 and the MS1 accuracy to 10. The unrelated runs, isotopologues, match-between-runs and no shared spectra options were enabled, whereas the heuristic inference was disabled. The protein inference was set to Genes and the IDs, RT and IM profiling was used for the library generation. The FDR was at the default 1%.

The data was searched initially against the reviewed Uniprot human protein database (canonical and isoforms) with ∼42000 entries (Release number 2023_09). The identified proteins were then merged with a previously described (85), in-house blood protein database, that consists of 1178 canonical and 1267 mutation sequences, to avoid false discoveries. The dengue virus II polyprotein sequence was also included in the database. The main report of DIA-NN (Q.Value, Lib.Q.Value and Lib.PG.Q.Value at 1%) was filtered from proteins with less than 2 identified peptides and methionine-oxidized peptides, and used to obtain batch-adjusted peptide intensities using the ComBat approach, based on the empirical Bayes methodology (87) in the sva package (88), as well as for the generation of label-free quantification (LFQ) intensities. Lastly, proteins describing the variable domain of immunoglobulins and contaminant plasma proteins downloaded from github (https://github.com/MannLabs/Quality-Control-of-the-Plasma-Proteome/blob/master/data/Marker%20List.xlsx), were omitted to ensure data quality. A minimum value imputation per protein was performed to facilitate statistical analysis. A list of all identified proteins used for the statistical analysis after excluding contaminants, can be found in Supplementary Data S1. Of note, throughout this work, we refer to the quantified proteins by their corresponding gene names except for the viral protein NS1. To distinguish AP from RP, the log2-transformed protein abundances (LFQ) were analyzed using linear mixed-effects models with phase, clinical classification, and their interaction as fixed effects, and the subjects as a random intercept. The filtered protein panel with adjusted p-value < 0.05 is shown in Supplementary Data S2. The models were fitted by maximum likelihood, and pairwise group comparisons within each classification were conducted using estimated marginal means with 95% confidence intervals. For comparing the subclinical with the hospitalized patients in AP and CP, we fitted linear models on the log2-LFQ values of each phase separately using clinical classification as a fixed effect and filtered the proteins with adjusted p-value < 0.05 (Supplementary Data S3). Similarly, a linear model was fitted in the severe AP patients to distinguish DF from DHF/DSS cases, and a cutoff of p-value < 0.01 was set (Supplementary Data S4).

### Developing patient classification models

Two complementary classification approaches were developed to distinguish between hospitalized and subclinical as well as acute and recovery phase patients using LFQ proteomic data: logistic regression with elastic-net regularization and random forest. In each case, the patient LFQ values were the predictors for the respective time-points, and the two separate classes (subclinical vs hospitalized, AP vs RP) were the response variables. Both the approaches followed identical preprocessing and evaluation methods: patient data was transformed into wide format, with patients as rows and genes as columns. Patient-level stratified train-test splitting (75–25) was performed to prevent data leakage. A multi-stage feature selection was performed including a) Variance filtering, b) Correlation filtering, c) Random forest importance selection to select the top 100 features and finally, d) Boruta selection. Class imbalances were addressed via minority class up sampling with shuffling. Features were standardized using training-set parameters and evaluation employed “Leave-one-patient-out cross-validation (LOPO-CV)” with independent feature selection per fold. The performance of the models in each case was evaluated using a) Receiver Operating Characteristic-Area Under The Curve (ROC-AUC), b) Optimal threshold using Youden’s J, c) Sensitivity, d) Specificity, e) Positive Predictive Value (PPV) and finally f) Negative Predictive Value (NPV). After pre-processing, the predictions by each classifier were performed as described below. Of these, the random forest showed superior performance over the other classifiers.

Importantly, a 21 gene panel for the hospitalized vs subclinical cohort and an 18 gene panel for the dengue fever (DF) vs the dengue hemorrhagic fever (DHF) (based on Figures 3 and 4 of Results respectively) were used to train, test and cross-validate both the classifiers. All genes were used for the Acute Phase vs the Recovery Phase classifiers (AP vs RP).

#### Logistic regression with elastic-net regularization

Elastic-net logistic regression (glmnet) was implemented with hyperparameter tuning: alpha (0-1, step 0.05) and lambda (10-fold CV per alpha). Lambda.1se (one standard error rule) was selected for regularization. Optimal alpha maximized CV-AUC at lambda.1se. Feature importance was assessed via non-zero coefficients, with magnitude and sign indicating association strength and direction.

#### Random forest

The random forest hyperparameters were tuned with 5-fold cross validation as follows: (ntree (100, 200, 300, 500), mtry (√p, logL(p), p/3), nodesize (1, 3, 5, 10), and maxnodes (10, 20, 50, unlimited). Thirty parameter combinations were evaluated; optimal parameters maximized CV-AUC. Variable importance was assessed via Mean Decrease in Gini impurity and Mean Decrease in Accuracy.

## Data availability

The mass spectrometry-based plasma proteomics data have been deposited on PRIDE under the dataset identifier PXD074467 (token: Fpvsrnt9GqaP). The codes used to develop the classifiers have been deposited in the git repository “Dengue-patient-classifiers” at the following link: https://github.com/UU-DENV/Dengue-patient-classifiers

## Supplementary information

The supplementary information consists of a supplementary information file that contains Supplementary Table S1 and Supplementary Figures S1-S7, as well as Supplementary Data S1-S4.

## Data Availability

The mass spectrometry-based plasma proteomics data have been deposited on PRIDE under the dataset identifier PXD074467 (token: Fpvsrnt9GqaP). The codes used to develop the classifiers have been deposited in the git repository Dengue-patient-classifiers at the following link: https://github.com/UU-DENV/Dengue-patient-classifiers

## Acknowledgements

This research was funded by the Dutch Research Council NWO Gravitation 2013 BOO, Institute for Chemical Immunology (ICI; 024.002.009), the European Research Council Executive Agency HORIZON ERC-2022-STG (FLAVIR; 101077640) to J.S and NIH U01 NIH-PICREID (1U01AI151758) to T.C. The laboratory of T.C. is additionally funded by the NIH (R01 AI175134, R01 AI137276), Horizon Europe (grant no 101137033), ANR (grant no AAPG2023), Wellcome Trust (grant no 311543/Z/24/Z). A.J.R.H. acknowledges support of the Netherlands Organization for Scientific Research (Spinoza Award SPI.2017.028).

We would like to thank Dr. Rithea Leang from the National Center for parasitology, entomology and Malaria Control, Ministry of Health, Cambodia, for facilitating the study. We would like to thank Borita Heng, Sivleangkalyaney Loek, Phalla Y, Leangyi Heng, Sreymom Ken and Sopheak Sorn for their help in patient inclusions, sample collection, processing, DENV diagnosis and patient characterization.

We would like to thank all patients and legal guardians who participated in the study. We thank the doctors and nurses at Baray-Santuk referral hospital, Stoung referral hospital, and Kampong Thom provincial hospital in Kampong Thom province and Jayavaraman VII Hospital in Siem Reap province for patient enrolment and sample collection.

## Declaration of possible conflicts of interests

All authors declare no competing interests.

## Author Contribution

Conceptualization: T.M.S., J.S.,T.C.

Methodology: T.M.S., S.K.N.

Formal Analysis: S.K.N., A.A.K.

Investigation: T.M.S., S.K.N., S.Lay.

Resources: J.S., A.J.R.H., V.D., S.Ly

Data Curation: S.K.N., A.A.K

Writing – Original Draft: T.M.S.

Writing – Review & Editing: T.M.S., S.K.N., A.A.K., J.S., A.J.R.H., S.Lay, T.C.

Visualization: S.K,N., A.A.K

Supervision: J.S., A.J.R.H, S.Ly, V.D., T.C.

Project Administration: T.C.

Funding Acquisition: J.S., A.J.R.H.,T.C.

## References

1. Paz-Bailey G, Adams LE, Deen J, Anderson KB, Katzelnick LC. Dengue. The Lancet. 2024 Feb;403(10427):667–82. doi:10.1016/S0140-6736(23)02576-X

2. Malavige GN, Ogg GS. Molecular mechanisms in the pathogenesis of dengue infections. Trends Mol Med. 2024 May;30(5):484–98. doi:10.1016/j.molmed.2024.03.006

3. Guzman MG, Halstead SB, Artsob H, Buchy P, Farrar J, Gubler DJ, et al. Dengue: a continuing global threat. Nat Rev Microbiol. 2010 Dec;8(12 Suppl):S7–16. doi:10.1038/nrmicro2460 PubMed PMID: 21079655; PubMed Central PMCID: PMC4333201.

4. Postler TS, Beer M, Blitvich BJ, Bukh J, De Lamballerie X, Drexler JF, et al. Renaming of the genus Flavivirus to Orthoflavivirus and extension of binomial species names within the family Flaviviridae. Arch Virol. 2023 Sep;168(9):224, s00705-023-05835–1. doi:10.1007/s00705-023-05835-1

5. Bhatt S, Gething PW, Brady OJ, Messina JP, Farlow AW, Moyes CL, et al. The global distribution and burden of dengue. Nature. 2013 Apr 25;496(7446):504–7. doi:10.1038/nature12060 PubMed PMID: 23563266; PubMed Central PMCID: PMC3651993.

6. Simmons CP, Farrar JJ, Nguyen van VC, Wills B. Dengue. N Engl J Med. 2012 Apr 12;366(15):1423–32. doi:10.1056/NEJMra1110265 PubMed PMID: 22494122.

7. Rodriguez-Barraquer I, Salje H, Cummings DA. Opportunities for improved surveillance and control of dengue from age-specific case data. eLife. 2019 May 23;8:e45474. doi:10.7554/eLife.45474

8. Deng J, Zhang H, Wang Y, Liu Q, Du M, Yan W, et al. Global, regional, and national burden of dengue infection in children and adolescents: an analysis of the Global Burden of Disease Study 2021. EClinicalMedicine. 2024 Dec;78:102943. doi:10.1016/j.eclinm.2024.102943 PubMed PMID: 39640938; PubMed Central PMCID: PMC11617407.

9. Verhagen LM, De Groot R. Dengue in children. J Infect. 2014 Nov;69:S77–86. doi:10.1016/j.jinf.2014.07.020

10. Malavige GN, Sjö P, Singh K, Piedagnel JM, Mowbray C, Estani S, et al. Facing the escalating burden of dengue: Challenges and perspectives. PLOS Glob Public Health. 2023;3(12):e0002598. doi:10.1371/journal.pgph.0002598 PubMed PMID: 38100392; PubMed Central PMCID: PMC10723676.

11. Thomas SJ. Is new dengue vaccine efficacy data a relief or cause for concern? Npj Vaccines. 2023 Apr 15;8(1):55. doi:10.1038/s41541-023-00658-2

12. Kalimuddin S, Chia PY, Low JG, Ooi EE. Dengue and severe dengue. Clin Microbiol Rev. 2025 Dec 11;38(4):e0024424. doi:10.1128/cmr.00244-24 PubMed PMID: 40910631; PubMed Central PMCID: PMC12697202.

13. Puerta-Guardo H, Glasner DR, Harris E. Dengue Virus NS1 Disrupts the Endothelial Glycocalyx, Leading to Hyperpermeability. PLoS Pathog. 2016 Jul;12(7):e1005738. doi:10.1371/journal.ppat.1005738 PubMed PMID: 27416066; PubMed Central PMCID: PMC4944995.

14. Rastogi M, Sharma N, Singh SK. Flavivirus NS1: a multifaceted enigmatic viral protein. Virol J. 2016 Dec;13(1):131. doi:10.1186/s12985-016-0590-7

15. Hadpech S, Thongboonkerd V. Proteomic investigations of dengue virus infection: key discoveries over the last 10 years. Expert Rev Proteomics. 2024;21(7–8):281–95. doi:10.1080/14789450.2024.2383580 PubMed PMID: 39049185.

16. Garishah FM, Boahen CK, Vadaq N, Pramudo SG, Tunjungputri RN, Riswari SF, et al. Longitudinal proteomic profiling of the inflammatory response in dengue patients. Rayner S, editor. PLoS Negl Trop Dis. 2023 Jan 3;17(1):e0011041. doi:10.1371/journal.pntd.0011041

17. Choudhury KR, Verma P, Ray AG, Samanta S, Manna A, Bandyopadhyay A, et al. Differential Proteomic Profiling at Different Phases of Dengue Infection: An Intricate Insight from Proteins to Pathogenesis. J Proteome Res. 2024 Sep 6;23(9):3731–45. doi:10.1021/acs.jproteome.3c00751

18. Nhi DM, Huy NT, Ohyama K, Kimura D, Lan NTP, Uchida L, et al. A Proteomic Approach Identifies Candidate Early Biomarkers to Predict Severe Dengue in Children. Halstead SB, editor. PLoS Negl Trop Dis. 2016 Feb 19;10(2):e0004435. doi:10.1371/journal.pntd.0004435

19. Kumar Y, Liang C, Bo Z, Rajapakse JC, Ooi EE, Tannenbaum SR. Serum Proteome and Cytokine Analysis in a Longitudinal Cohort of Adults with Primary Dengue Infection Reveals Predictive Markers of DHF. Rothman AL, editor. PLoS Negl Trop Dis. 2012 Nov 29;6(11):e1887. doi:10.1371/journal.pntd.0001887

20. Ray S, Srivastava R, Tripathi K, Vaibhav V, Patankar S, Srivastava S. Serum Proteome Changes in Dengue Virus-Infected Patients from a Dengue-Endemic Area of India: Towards New Molecular Targets? OMICS J Integr Biol. 2012 Oct;16(10):527–36. doi:10.1089/omi.2012.0037

21. Han L, Ao X, Lin S, Guan S, Zheng L, Han X, et al. Quantitative Comparative Proteomics Reveal Biomarkers for Dengue Disease Severity. Front Microbiol. 2019 Dec 10;10:2836. doi:10.3389/fmicb.2019.02836

22. Fragnoud R, Flamand M, Reynier F, Buchy P, Duong V, Pachot A, et al. Differential proteomic analysis of virus-enriched fractions obtained from plasma pools of patients with dengue fever or severe dengue. BMC Infect Dis. 2015 Dec;15(1):518. doi:10.1186/s12879-015-1271-7

23. Chumchanchira C, Ramphan S, Paemanee A, Roytrakul S, Lithanatudom P, Smith DR. A 2D-proteomic analysis identifies proteins differentially regulated by two different dengue virus serotypes. Sci Rep. 2024 Apr 9;14(1):8287. doi:10.1038/s41598-024-57930-1 PubMed PMID: 38594317; PubMed Central PMCID: PMC11003990.

24. Organization WH. Dengue haemorrhagic fever: diagnosis, treatment, prevention and control. World Health Organization; 1997.

25. Geyer PE, Voytik E, Treit PV, Doll S, Kleinhempel A, Niu L, et al. Plasma Proteome Profiling to detect and avoid sample-related biases in biomarker studies. EMBO Mol Med. 2019 Nov 7;11(11):e10427. doi:10.15252/emmm.201910427

26. Chien YW, Chuang HN, Wang YP, Perng GC, Chi CY, Shih HI. Short-term, medium-term, and long-term risks of nonvariceal upper gastrointestinal bleeding after dengue virus infection. Ribeiro GS, editor. PLoS Negl Trop Dis. 2022 Jan 19;16(1):e0010039. doi:10.1371/journal.pntd.0010039

27. Kalaidopoulou Nteak S, Völlmy F, Lukassen MV, Van Den Toorn H, Den Boer MA, Bondt A, et al. Longitudinal Fluctuations in Protein Concentrations and Higher-Order Structures in the Plasma Proteome of Kidney Failure Patients Subjected to a Kidney Transplant. J Proteome Res. 2024 Jun 7;23(6):2124–36. doi:10.1021/acs.jproteome.4c00064

28. Völlmy F, van den Toorn H, Zenezini Chiozzi R, Zucchetti O, Papi A, Volta CA, et al. A serum proteome signature to predict mortality in severe COVID-19 patients. Life Sci Alliance. 2021 Sep;4(9):e202101099. doi:10.26508/lsa.202101099 PubMed PMID: 34226277; PubMed Central PMCID: PMC8321673.

29. Yu Z, Xiao C, Liu R, Pi D, Jin B, Zou Z, et al. The protective effect of apolipoprotein H in paediatric sepsis. Crit Care Lond Engl. 2024 Jan 30;28(1):36. doi:10.1186/s13054-024-04809-2 PubMed PMID: 38291524; PubMed Central PMCID: PMC10826270.

30. Garcinuño S, Lalueza A, Gil-Etayo FJ, Díaz-Simón R, Lizasoain I, Moraga A, et al. Immune dysregulation is an important factor in the underlying complications in Influenza infection. ApoH, IL-8 and IL-15 as markers of prognosis. Front Immunol. 2024;15:1443096. doi:10.3389/fimmu.2024.1443096 PubMed PMID: 39176097; PubMed Central PMCID: PMC11339618.

31. Hu L, Yao W, Wang F, Rong X, Peng T. GP73 is upregulated by hepatitis C virus (HCV) infection and enhances HCV secretion. PloS One. 2014;9(3):e90553. doi:10.1371/journal.pone.0090553 PubMed PMID: 24608522; PubMed Central PMCID: PMC3946557.

32. Mantovani A, Garlanda C. Humoral Innate Immunity and Acute-Phase Proteins. N Engl J Med. 2023 Feb 2;388(5):439–52. doi:10.1056/NEJMra2206346 PubMed PMID: 36724330; PubMed Central PMCID: PMC9912245.

33. Stober VP, Lim YP, Opal S, Zhuo L, Kimata K, Garantziotis S. Inter-α-inhibitor Ameliorates Endothelial Inflammation in Sepsis. Lung. 2019 Jun;197(3):361–9. doi:10.1007/s00408-019-00228-1

34. Ulm C, Saffarzadeh M, Mahavadi P, Müller S, Prem G, Saboor F, et al. Soluble polysialylated NCAM: a novel player of the innate immune system in the lung. Cell Mol Life Sci CMLS. 2013 Oct;70(19):3695–708. doi:10.1007/s00018-013-1342-0 PubMed PMID: 23619613; PubMed Central PMCID: PMC11113884.

35. Liu KT, Liu YH, Chen YH, Lin CY, Huang CH, Yen MC, et al. Serum Galectin-9 and Galectin-3-Binding Protein in Acute Dengue Virus Infection. Int J Mol Sci. 2016 May 27;17(6):832. doi:10.3390/ijms17060832

36. Kostadinova L, Shive CL, Judge C, Zebrowski E, Compan A, Rife K, et al. During Hepatitis C Virus (HCV) Infection and HCV-HIV Coinfection, an Elevated Plasma Level of Autotaxin Is Associated With Lysophosphatidic Acid and Markers of Immune Activation That Normalize During Interferon-Free HCV Therapy. J Infect Dis. 2016 Nov 1;214(9):1438–48. doi:10.1093/infdis/jiw372 PubMed PMID: 27540113; PubMed Central PMCID: PMC6281372.

37. Kim YT, Huh JW, Choi YH, Yoon HK, Nguyen TT, Chun E, et al. Highly secreted tryptophanyl tRNA synthetase 1 as a potential theranostic target for hypercytokinemic severe sepsis. EMBO Mol Med. 2024 Jan;16(1):40–63. doi:10.1038/s44321-023-00004-y PubMed PMID: 38177528; PubMed Central PMCID: PMC10883277.

38. Yong YK, Tan HY, Jen SH, Shankar EM, Natkunam SK, Sathar J, et al. Aberrant monocyte responses predict and characterize dengue virus infection in individuals with severe disease. J Transl Med. 2017 May 31;15(1):121. doi:10.1186/s12967-017-1226-4 PubMed PMID: 28569153; PubMed Central PMCID: PMC5452397.

39. Bowers JR, Readler JM, Sharma P, Excoffon KJDA. Poliovirus Receptor: More than a simple viral receptor. Virus Res. 2017 Oct 15;242:1–6. doi:10.1016/j.virusres.2017.09.001 PubMed PMID: 28870470; PubMed Central PMCID: PMC5650920.

40. Dussart P, Duong V, Bleakley K, Fortas C, Lorn Try P, Kim KS, et al. Comparison of dengue case classification schemes and evaluation of biological changes in different dengue clinical patterns in a longitudinal follow-up of hospitalized children in Cambodia. Althouse B, editor. PLoS Negl Trop Dis. 2020 Sep 14;14(9):e0008603. doi:10.1371/journal.pntd.0008603

41. Upasani V, Ter Ellen BM, Sann S, Lay S, Heng S, Laurent D, et al. Characterization of soluble TLR2 and CD14 levels during acute dengue virus infection. Heliyon. 2023 Jun;9(6):e17265. doi:10.1016/j.heliyon.2023.e17265 PubMed PMID: 37416678; PubMed Central PMCID: PMC10320027.

42. Benfrid S, Park K, Dellarole M, Voss JE, Tamietti C, Pehau-Arnaudet G, et al. Dengue virus NS1 protein conveys pro-inflammatory signals by docking onto high-density lipoproteins. EMBO Rep. 2022 Jul 5;23(7):e53600. doi:10.15252/embr.202153600

43. Xu G, Xia Z, Deng F, Liu L, Wang Q, Yu Y, et al. Inducible LGALS3BP/90K activates antiviral innate immune responses by targeting TRAF6 and TRAF3 complex. PLoS Pathog. 2019 Aug;15(8):e1008002. doi:10.1371/journal.ppat.1008002 PubMed PMID: 31404116; PubMed Central PMCID: PMC6705879.

44. Ng WH, Liu X, Ling ZL, Santos CNO, Magalhães LS, Kueh AJ, et al. FHL1 promotes chikungunya and o’nyong-nyong virus infection and pathogenesis with implications for alphavirus vaccine design. Nat Commun. 2023 Oct 26;14(1):6605. doi:10.1038/s41467-023-42330-2 PubMed PMID: 37884534; PubMed Central PMCID: PMC10603155.

45. Dong J, Li S, Lu Z, Du P, Liu G, Li M, et al. HCMV-miR-US33-5p promotes apoptosis of aortic vascular smooth muscle cells by targeting EPAS1/SLC3A2 pathway. Cell Mol Biol Lett. 2022 Dec;27(1):40. doi:10.1186/s11658-022-00340-w

46. Shu T, Ning W, Wu D, Xu J, Han Q, Huang M, et al. Plasma Proteomics Identify Biomarkers and Pathogenesis of COVID-19. Immunity. 2020 Nov 17;53(5):1108–1122.e5. doi:10.1016/j.immuni.2020.10.008 PubMed PMID: 33128875; PubMed Central PMCID: PMC7574896.

47. Nikitopoulou I, Fanidis D, Ntatsoulis K, Moulos P, Mpekoulis G, Evangelidou M, et al. Increased Autotaxin Levels in Severe COVID-19, Correlating with IL-6 Levels, Endothelial Dysfunction Biomarkers, and Impaired Functions of Dendritic Cells. Int J Mol Sci. 2021 Sep 16;22(18):10006. doi:10.3390/ijms221810006 PubMed PMID: 34576169; PubMed Central PMCID: PMC8469279.

48. Carr JM, Cabezas-Falcon S, Dubowsky JG, Hulme-Jones J, Gordon DL. Dengue virus and the complement alternative pathway. FEBS Lett. 2020 Aug;594(16):2543–55. doi:10.1002/1873-3468.13730 PubMed PMID: 31943152.

49. Aviner R, Frydman J. Proteostasis in Viral Infection: Unfolding the Complex Virus-Chaperone Interplay. Cold Spring Harb Perspect Biol. 2020 Mar 2;12(3):a034090. doi:10.1101/cshperspect.a034090 PubMed PMID: 30858229; PubMed Central PMCID: PMC7050591.

50. Hafirassou ML, Meertens L, Umaña-Diaz C, Labeau A, Dejarnac O, Bonnet-Madin L, et al. A Global Interactome Map of the Dengue Virus NS1 Identifies Virus Restriction and Dependency Host Factors. Cell Rep. 2017 Dec;21(13):3900–13. doi:10.1016/j.celrep.2017.11.094

51. Swaine T, Dittmar MT. CDC42 Use in Viral Cell Entry Processes by RNA Viruses. Viruses. 2015 Dec 10;7(12):6526–36. doi:10.3390/v7122955 PubMed PMID: 26690467; PubMed Central PMCID: PMC4690878.

52. McCarthy MK, Weinberg JB. The immunoproteasome and viral infection: a complex regulator of inflammation. Front Microbiol. 2015 Jan 29;6. doi:10.3389/fmicb.2015.00021

53. Diosa-Toro M, Prasanth KR, Bradrick SS, Garcia Blanco MA. Role of RNA-binding proteins during the late stages of Flavivirus replication cycle. Virol J. 2020 Apr 25;17(1):60. doi:10.1186/s12985-020-01329-7 PubMed PMID: 32334603; PubMed Central PMCID: PMC7183730.

54. Lopez-Nieto M, Locker N. Understanding the mechanisms of mitochondrial rewiring during viral infections. J Gen Virol. 2025 Jul;106(7):002128. doi:10.1099/jgv.0.002128 PubMed PMID: 40622847; PubMed Central PMCID: PMC12282254.

55. Sheng Y, Deng Y, Li X, Ji P, Sun X, Liu B, et al. Hepatitis E virus ORF3 protein hijacking thioredoxin domain-containing protein 5 (TXNDC5) for its stability to promote viral particle release. Ou JHJ, editor. J Virol. 2024 Apr 16;98(4):e01649–23. doi:10.1128/jvi.01649-23

56. Clarke MCH, Talib S, Figg NL, Bennett MR. Vascular Smooth Muscle Cell Apoptosis Induces Interleukin-1–Directed Inflammation: Effects of Hyperlipidemia-Mediated Inhibition of Phagocytosis. Circ Res. 2010 Feb 5;106(2):363–72. doi:10.1161/CIRCRESAHA.109.208389

57. Arreola-Diaz R, Majluf-Cruz A, Sanchez-Torres L, Hernandez-Juarez J. The Pathophysiology of The Antiphospholipid Syndrome: A Perspective From The Blood Coagulation System. Clin Appl Thromb. 2022 Jan;28:10760296221088576. doi:10.1177/10760296221088576

58. Pober JS. Endothelial activation: intracellular signaling pathways. Arthritis Res. 2002;4 Suppl 3(Suppl 3):S109–116. doi:10.1186/ar576 PubMed PMID: 12110129; PubMed Central PMCID: PMC3240152.

59. Osborn L, Hession C, Tizard R, Vassallo C, Luhowskyj S, Chi-Rosso G, et al. Direct expression cloning of vascular cell adhesion molecule 1, a cytokine-induced endothelial protein that binds to lymphocytes. Cell. 1989 Dec 22;59(6):1203–11. doi:10.1016/0092-8674(89)90775-7

60. van de Weg CAM, Koraka P, van Gorp ECM, Mairuhu ATA, Supriatna M, Soemantri A, et al. Lipopolysaccharide levels are elevated in dengue virus infected patients and correlate with disease severity. J Clin Virol Off Publ Pan Am Soc Clin Virol. 2012 Jan;53(1):38–42. doi:10.1016/j.jcv.2011.09.028 PubMed PMID: 22014848.

61. Wight TN, Kang I, Evanko SP, Harten IA, Chang MY, Pearce OMT, et al. Versican-A Critical Extracellular Matrix Regulator of Immunity and Inflammation. Front Immunol. 2020;11:512. doi:10.3389/fimmu.2020.00512 PubMed PMID: 32265939; PubMed Central PMCID: PMC7105702.

62. Chanh HQ, Trieu HT, Tran Kim H, Huynh Ngoc Thien V, Huyen VNT, Moncada A, et al. Kinetics of cardiovascular and inflammatory biomarkers in paediatric dengue shock syndrome. Oxf Open Immunol. 2024;5(1):iqae005. doi:10.1093/oxfimm/iqae005 PubMed PMID: 39193474; PubMed Central PMCID: PMC11211616.

63. Escalante NK, von Rossum A, Lee M, Choy JC. CD155 on human vascular endothelial cells attenuates the acquisition of effector functions in CD8 T cells. Arterioscler Thromb Vasc Biol. 2011 May;31(5):1177–84. doi:10.1161/ATVBAHA.111.224162 PubMed PMID: 21330602.

64. Luo Z, Lei H, Sun Y, Liu X, Su DF. Orosomucoid, an acute response protein with multiple modulating activities. J Physiol Biochem. 2015 Jun;71(2):329–40. doi:10.1007/s13105-015-0389-9 PubMed PMID: 25711902.

65. Cabezas-Falcon S, Norbury AJ, Hulme-Jones J, Klebe S, Adamson P, Rudd PA, et al. Changes in complement alternative pathway components, factor B and factor H during dengue virus infection in the AG129 mouse. J Gen Virol. 2021 Mar;102(3):001547. doi:10.1099/jgv.0.001547 PubMed PMID: 33410734; PubMed Central PMCID: PMC8515863.

66. Dobó J, Kocsis A, Farkas B, Demeter F, Cervenak L, Gál P. The Lectin Pathway of the Complement System-Activation, Regulation, Disease Connections and Interplay with Other (Proteolytic) Systems. Int J Mol Sci. 2024 Jan 26;25(3):1566. doi:10.3390/ijms25031566 PubMed PMID: 38338844; PubMed Central PMCID: PMC10855846.

67. Byrne AB, Talarico LB. Role of the complement system in antibody-dependent enhancement of flavivirus infections. Int J Infect Dis. 2021 Feb;103:404–11. doi:10.1016/j.ijid.2020.12.039

68. Gan ES, Tan HC, Le DHT, Huynh TT, Wills B, Seidah NG, et al. Dengue virus induces PCSK9 expression to alter antiviral responses and disease outcomes. J Clin Invest. 2020 Aug 31;130(10):5223–34. doi:10.1172/JCI137536

69. Morley SC. The actin-bundling protein L-plastin: a critical regulator of immune cell function. Int J Cell Biol. 2012;2012:935173. doi:10.1155/2012/935173 PubMed PMID: 22194750; PubMed Central PMCID: PMC3238366.

70. Viettri M, Zambrano JL, Rosales R, Caraballo GI, Gutiérrez-Escolano AL, Ludert JE. Flavivirus infections induce a Golgi stress response in vertebrate and mosquito cells. Sci Rep. 2021 Dec 6;11(1):23489. doi:10.1038/s41598-021-02929-1 PubMed PMID: 34873243; PubMed Central PMCID: PMC8648732.

71. Brunetta E, Folci M, Bottazzi B, De Santis M, Gritti G, Protti A, et al. Macrophage expression and prognostic significance of the long pentraxin PTX3 in COVID-19. Nat Immunol. 2021 Jan;22(1):19–24. doi:10.1038/s41590-020-00832-x PubMed PMID: 33208929.

72. Gutmann C, Takov K, Burnap SA, Singh B, Ali H, Theofilatos K, et al. SARS-CoV-2 RNAemia and proteomic trajectories inform prognostication in COVID-19 patients admitted to intensive care. Nat Commun. 2021 Jun 7;12(1):3406. doi:10.1038/s41467-021-23494-1 PubMed PMID: 34099652; PubMed Central PMCID: PMC8184784.

73. Mairuhu ATA, Peri G, Setiati TE, Hack CE, Koraka P, Soemantri A, et al. Elevated plasma levels of the long pentraxin, pentraxin 3, in severe dengue virus infections. J Med Virol. 2005 Aug;76(4):547–52. doi:10.1002/jmv.20397 PubMed PMID: 15977234.

74. Samarasingha P, Karunatilake H, Jayanaga A, Jayawardhana H, Priyankara D. Dengue rhabdomyolysis successfully treated with hemoperfusion using CytoSorb® in combination with continuous renal replacement therapy: a case report. J Med Case Reports. 2024 Jul 19;18(1):329. doi:10.1186/s13256-024-04661-6 PubMed PMID: 39026342; PubMed Central PMCID: PMC11264817.

75. Md Sani SS, Han WH, Bujang MA, Ding HJ, Ng KL, Amir Shariffuddin MA. Evaluation of creatine kinase and liver enzymes in identification of severe dengue. BMC Infect Dis. 2017 Jul 21;17(1):505. doi:10.1186/s12879-017-2601-8 PubMed PMID: 28732476; PubMed Central PMCID: PMC5520296.

76. Pierce-Ruiz C, Santana WI, Sutton WJH, Fischler DA, Cooper HC, Marc LR, et al. Quantification of SARS-CoV-2 spike and nucleocapsid proteins using isotope dilution tandem mass spectrometry. Vaccine. 2021 Aug 23;39(36):5106–15. doi:10.1016/j.vaccine.2021.07.066 PubMed PMID: 34344552; PubMed Central PMCID: PMC8302847.

77. Drouin N, Elfrink HL, Boers SA, Van Hugten S, Wessels E, De Vries JJC, et al. A Targeted LC-MRM3 Proteomic Approach for the Diagnosis of SARS-CoV-2 Infection in Nasopharyngeal Swabs. Mol Cell Proteomics. 2024 Jul;23(7):100805. doi:10.1016/j.mcpro.2024.100805

78. Huang C, Hu X, Wang D, Gong R, Wang Q, Ren F, et al. Multi-cohort study on cytokine and chemokine profiles in the progression of COVID-19. Sci Rep. 2024 May 6;14(1):10324. doi:10.1038/s41598-024-61133-z

79. Yue R, Shen B, Morrison SJ. Clec11a/osteolectin is an osteogenic growth factor that promotes the maintenance of the adult skeleton. eLife. 2016 Dec 13;5:e18782. doi:10.7554/eLife.18782 PubMed PMID: 27976999; PubMed Central PMCID: PMC5158134.

80. Cardona-Ospina JA, Roy V, Marcano-Jiménez DE, Bos S, Duarte E, Zambrana JV, et al. IgA-driven neutrophil activation underlies severe dengue disease after primary Zika virus infection in humans. Nat Immunol. 2026 Jan;27(1):126–34. doi:10.1038/s41590-025-02363-9 PubMed PMID: 41482554.

81. Wegman AD, Waldran MJ, Bahr LE, Lu JQ, Baxter KE, Thomas SJ, et al. DENV-specific IgA contributes protective and non-pathologic function during antibody-dependent enhancement of DENV infection. PLoS Pathog. 2023 Aug;19(8):e1011616. doi:10.1371/journal.ppat.1011616 PubMed PMID: 37639455; PubMed Central PMCID: PMC10491401.

82. Ou TP, Yun C, Auerswald H, In S, Leang R, Huy R, et al. Improved detection of dengue and Zika viruses using multiplex RT-qPCR assays. J Virol Methods. 2020 Aug;282:113862. doi:10.1016/j.jviromet.2020.113862 PubMed PMID: 32417207.

83. World Health Organization (1997) Dengue haemorrhagic fever: diagnosis, treatment, prevention and control. 2nd edition.

84. Dengue: Guidelines for Diagnosis, Treatment, Prevention and Control: New Edition [Internet]. Geneva: World Health Organization; 2009 [cited 2026 Jan 23]. (WHO Guidelines Approved by the Guidelines Review Committee). Available from: http://www.ncbi.nlm.nih.gov/books/NBK143157/ PubMed PMID: 23762963.

85. Michalik S, Kalaidopoulou Nteak S, Drouin N, Salazar MG, Hammer E, Dhople VM, et al. Immunoglobulin sub-class levels define inter-donor plasma variability: a longitudinal dual-lab study [Internet]. 2025 [cited 2025 Dec 7]. Available from: http://biorxiv.org/lookup/doi/10.1101/2025.11.04.686385 doi:10.1101/2025.11.04.686385

86. Demichev V, Messner CB, Vernardis SI, Lilley KS, Ralser M. DIA-NN: neural networks and interference correction enable deep proteome coverage in high throughput. Nat Methods. 2020 Jan;17(1):41–4. doi:10.1038/s41592-019-0638-x PubMed PMID: 31768060; PubMed Central PMCID: PMC6949130.

87. Johnson WE, Li C, Rabinovic A. Adjusting batch effects in microarray expression data using empirical Bayes methods. Biostatistics. 2007 Jan 1;8(1):118–27. doi:10.1093/biostatistics/kxj037

88. Leek JT, Johnson WE, Parker HS, Jaffe AE, Storey JD. The sva package for removing batch effects and other unwanted variation in high-throughput experiments. Bioinformatics. 2012 Mar 15;28(6):882–3. doi:10.1093/bioinformatics/bts034

