## Supplementary Information for "Plasma proteomics identifies early markers of endothelial and inflammatory activation associated with dengue disease severity in children"

### Table of Contents


| | Page |
|---|---|
| **Supplementary Table S1 \|** Demographic and clinical characteristics of patients and healthy donors | **S3** |
| **Supplementary Figure S1 \|** Logistic regression classifier results discriminating disease severity in the acute and recovery phases of dengue disease. | **S4** |
| **Supplementary Figure S2 \|** Detection of NS1 structural protein of dengue virus in patient plasma. | **S6** |
| **Supplementary Figure S3 \|** Longitudinal profiles of immunoglobulins (IgG subclasses, IgA subclasses, IgM, and IgD) in subclinical and hospitalized patients across all dengue disease phases. | **S7** |
| **Supplementary Figure S4 \|** Violin plots of immunoglobulins (IgG, all subclasses, IgA, all subclasses, IgM, and IgD) comparing subclinical, hospitalized, and healthy individuals during the acute phase of the dengue disease | **S8** |
| **Supplementary Figure S5 \|** Violin plots of immunoglobulins (IgG, all subclasses, IgA, all subclasses, IgM, and IgD) comparing subclinical, hospitalized, and healthy individuals during the critical phase of the dengue disease | **S9** |
| **Supplementary Figure S6 \|** Violin plots of immunoglobulins (IgG, all subclasses, IgA, all subclasses, IgM, and IgD) comparing subclinical, hospitalized, and healthy individuals during the early recovery phase of the dengue disease | **S10** |
| **Supplementary Figure S7 \|** Violin plots of immunoglobulins (IgG, all subclasses, IgA, all subclasses, IgM, and IgD) comparing subclinical, hospitalized, and healthy individuals during the recovery phase of the dengue disease | **S11** |
| **Supplementary Data S1 \|** Quantitative overview of the relative abundance of the ~500 most abundant proteins across all samples | **xlsx** |
| **Supplementary Data S2 \|** Significantly differentiated proteins between acute and critical phases | **xlsx** |
| **Supplementary Data S3 \|** Significantly differentiated proteins between subclinical and hospitalized patients in acute and critical phases | **xlsx** |
| **Supplementary Data S4 \|** Significantly differentiated proteins between DF and DHF/DSS patients in acute phase | **xlsx** |

***Supplementary Table 1.*** *Demographic and clinical characteristics of patients and healthy donors. All Dengue cases were undergoing RT-PCR confirmed DENV2 infection and secondary infection as determined by HIA test on D0 and D10 plasma samples.*

| **Cohort 1** | HD (N=10) | Subclinical Dengue (N=23) | DF (N=43) | DHF/DSS (N=26) | Overall (N=102) |
|---|---|---|---|---|---|
| **Age (year)** | | | | | |
| Mean (SD) | 4.3 (3.3) | 16.9 (15.5) | 13.7 (7.6) | 11 (4.1) | 13.7 (9.7) |
| **Gender** | | | | | |
| F (%) | 6 (60%) | 10 (43%) | 18 (42%) | 15 (58%) | 49 (48%) |
| M (%) | 4 (40%) | 13 (57%) | 25 (58%) | 11 (42%) | 53 (52%) |
| **Day of fever** | | | | | |
| Mean (SD) | NA | 2.7 (2.9) | 2.2 (1.0) | 2.7 (1.5) | 2.4 (1.6) |
| **RT-qPCR (Ct value)** | | | | | |
| Mean (SD) | NA | 31.0 (6.4) | 25.6 (6.6) | 31.8 (5.5) | 28.7 (6.8) |
| **No of samples in each phase of infection** | | | | | |
| Acute phase (AP) | NA | 15 | 40 | 18 | 73 |
| Critical phase (CP) | NA | 8 | 41 | 19 | 68 |
| Early recovery phase (EP) | NA | 19 | 40 | 20 | 79 |
| Recovery phase (RP) | NA | 6 | 40 | 20 | 66 |

HD: healthy donor, DF: dengue fever, DHF: dengue hemorrhagic fever, DSS: dengue shock syndrome, SD: standard deviation, DENV2: dengue virus serotype 2, RT-qPCR: Reverse Transcriptase quantitative Polymerase Chain Reaction.

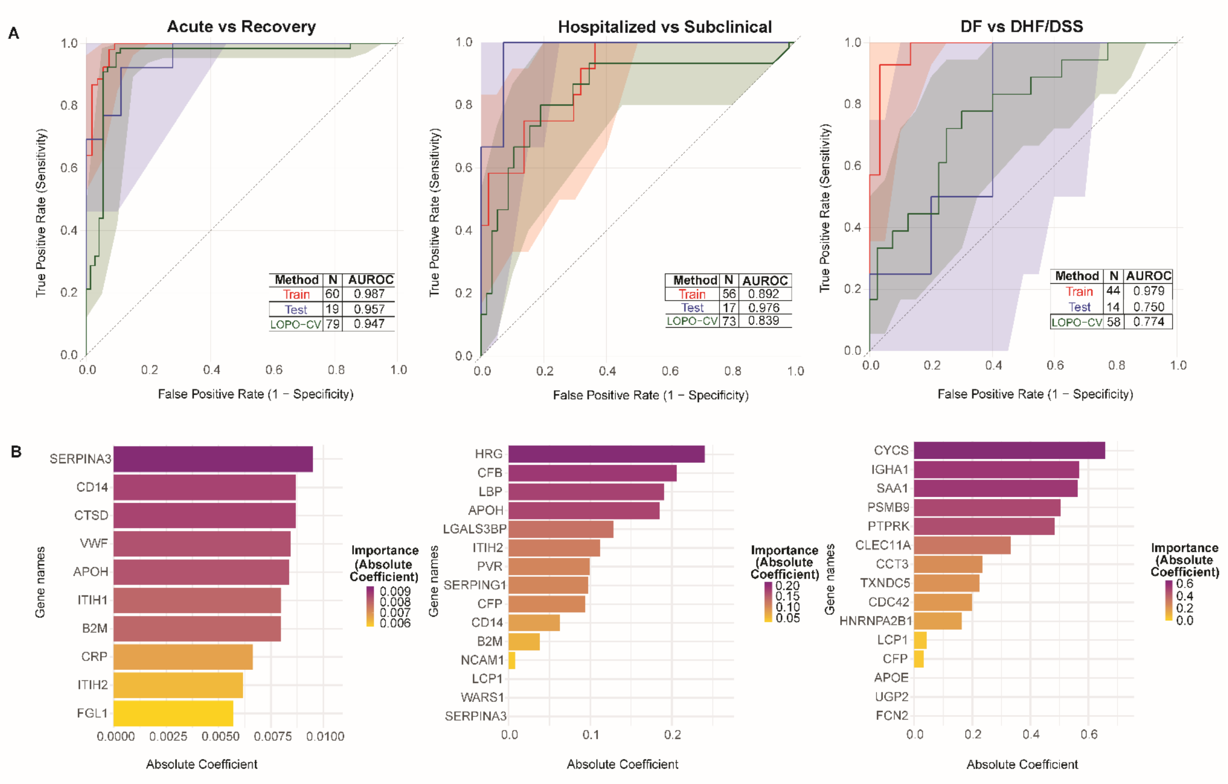


***Supplementary Figure S1**: Logistic regression classifier results discriminating disease severity in the acute and recovery phases of dengue disease.*

*A) Performance metrics of logistic regression classifiers distinguishing acute versus recovery phases (left), hospitalized versus subclinical cases (center), and dengue fever versus dengue hemorrhagic fever (right). The logistic regression models achieve strong discrimination across all comparisons based on their Leave-One-Patient-Out Cross-Validation (LOPO-CV) Area Under the Receiver Operating Characteristic Curve (AUROC) values: acute versus recovery phases (AUROC = 0.947, left), hospitalized versus subclinical (AUROC = 0.839, center), and dengue fever versus dengue hemorrhagic fever (AUROC = 0.774, right). Test set performance shows even higher discrimination, with AUROC values of 0.987, 0.876, and 0.979, respectively. All models substantially outperform the diagonal reference line representing random classification (AUROC = 0.5). Shaded regions indicate 95% confidence intervals for each ROC curve. B) The top 15 features ranked by absolute coefficient value from each model. Absolute coefficient measures the magnitude of influence each variable has on the log-odds of classification; higher values indicate greater discriminative importance. The most discriminative genes are SERPINA3 and CD14 for acute versus recovery phase classification, HRG and CFB for hospitalized versus subclinical classification, and CYCS and IGHA1 for dengue fever versus dengue hemorrhagic fever classification*.

A

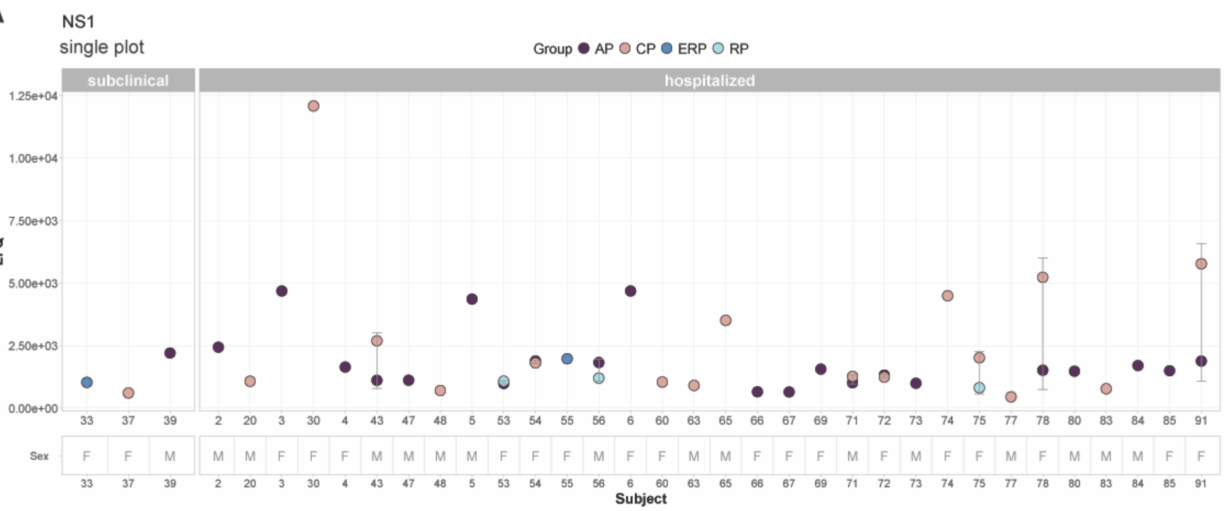


B

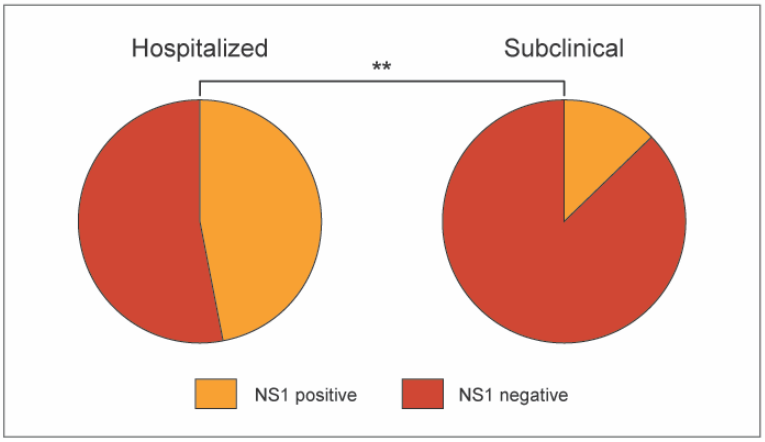


***Supplementary Figure S2. Detection of NS1 structural protein of dengue virus in patient plasma.***

*A) Left side of the plot represents the subclinical cases and the right side of the plot represents the hospitalized patients. The values on the y-axes represent the abundance of the protein in plasma in the form of label free quantitation (LFQ) intensity. For each subject the LFQ intensity of the NS1 protein in each available group (AP, CP, ERP, RP) is depicted and the error bars are showing the difference between the two groups. B) Pie charts representing percentage of NS1+ patients in each patient group. P-value is calculated using Fisher's exact test.*

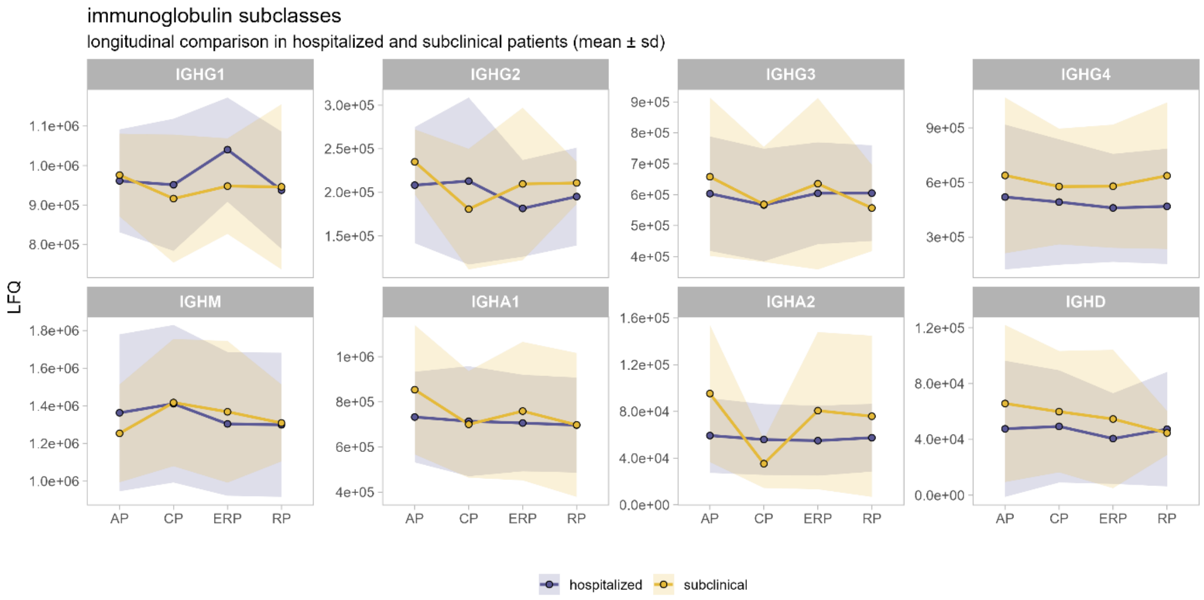


***Supplementary Figure S3. Longitudinal profiles of immunoglobulins (IgG subclasses, IgA subclasses, IgM, and IgD) in subclinical and hospitalized patients across all dengue disease phases.***

*The values on the y-axes represent the abundance of the protein in plasma in the form of label free quantitation (LFQ) intensity. The solid lines show the average protein abundance (LFQ) of each protein in each phase, and the faded purple or yellow areas indicate the standard deviation of each phase in the hospitalized patients or subclinical cases, respectively.*

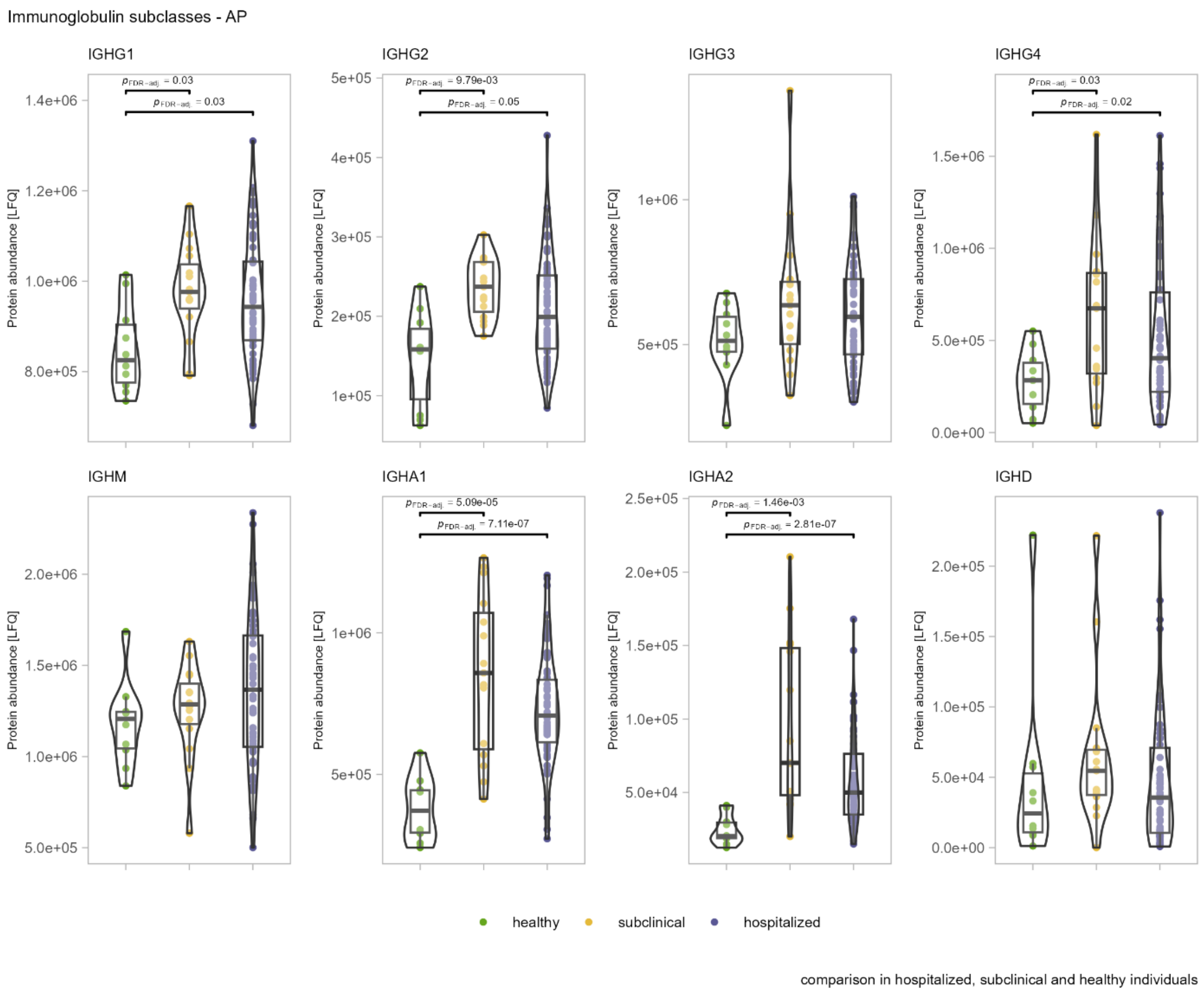


***Supplementary Figure S4. Violin plots of immunoglobulins (IgG, all subclasses, IgA, all subclasses, IgM, and IgD) comparing subclinical, hospitalized, and healthy individuals during the acute phase of the dengue disease.***

*The values on the y-axes represent the abundance of the protein in plasma in the form of label free quantitation (LFQ) intensity. Green dot violin represents healthy donors (left); yellow dot violin represents subclinical cases (center); purple dot violin represents hospitalized patients (right). The p-values of statistical significance are shown above the respective violins.*

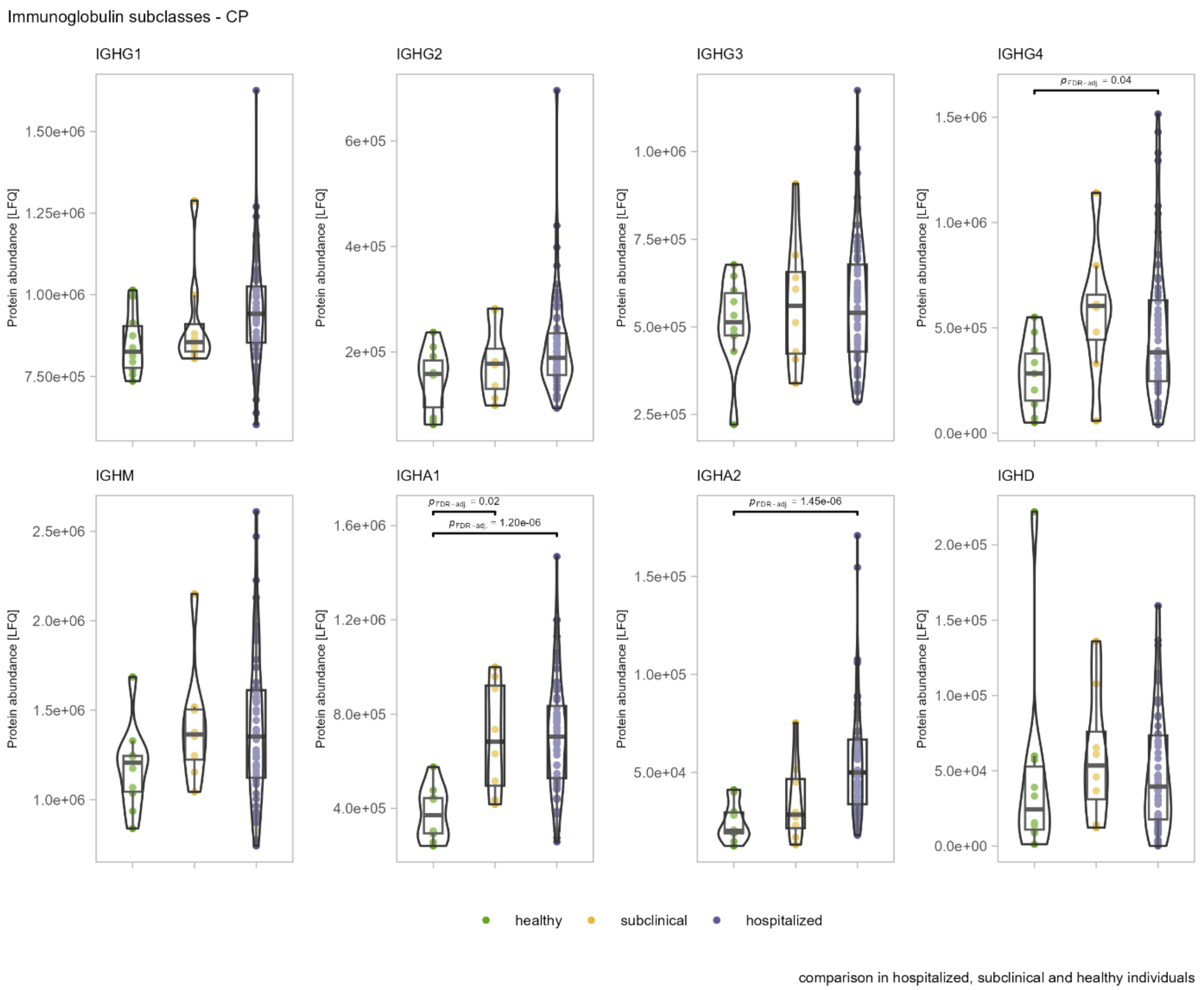


***Supplementary Figure S5. Violin plots of immunoglobulins (IgG subclasses, IgA subclasses, IgM, and IgD) comparing subclinical, hospitalized, and healthy individuals during the critical phase of the dengue disease.***

*The values on the y-axes represent the abundance of the protein in plasma in the form of label free quantitation (LFQ) intensity. Green dot violin represents healthy donors (left); yellow dot violin represents subclinical cases (center); purple dot violin represents hospitalized patients (right). The p-values of statistical significance are shown above the respective violins.*

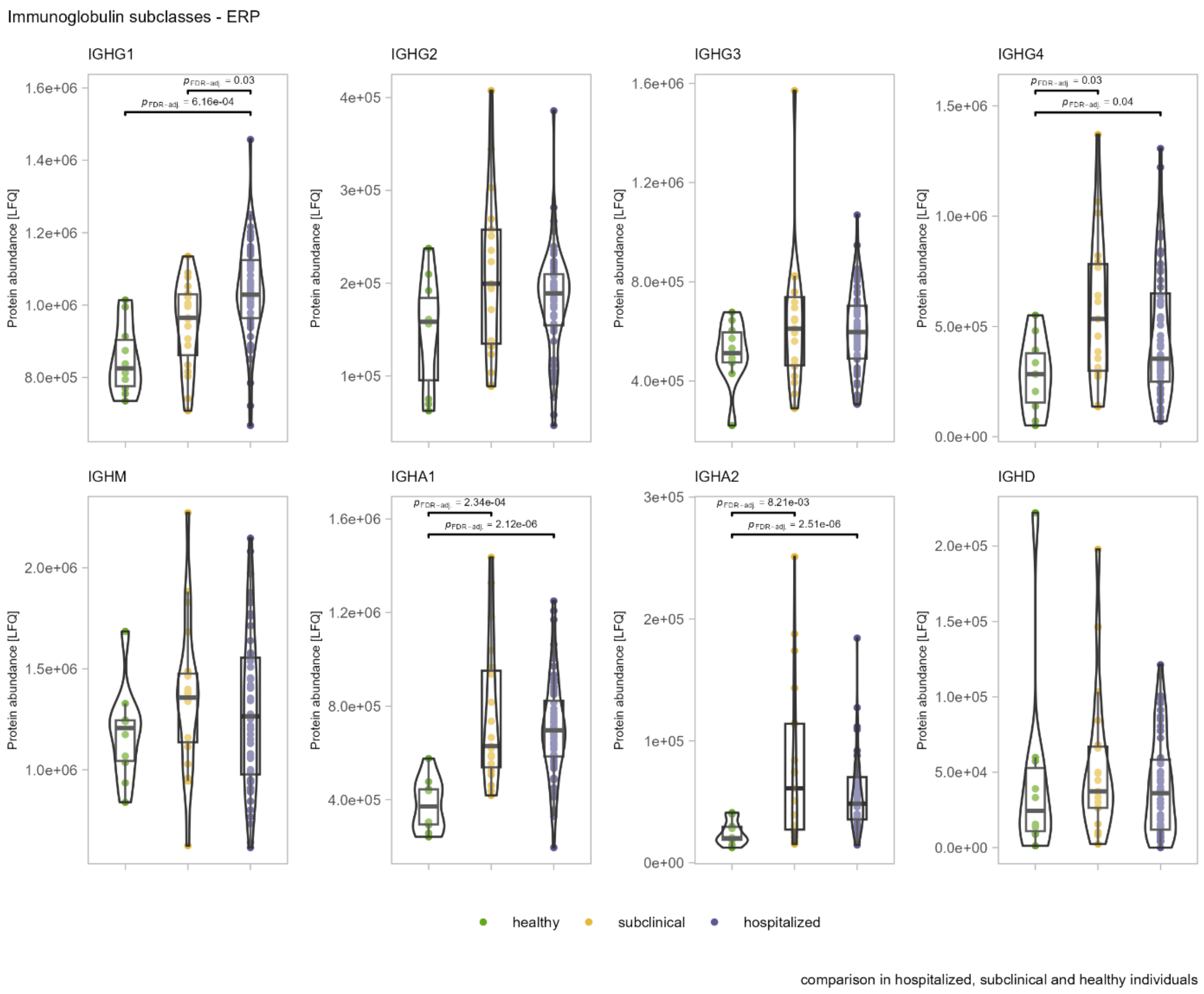


***Supplementary Figure S6. Violin plots of immunoglobulins (IgG subclasses, IgA subclasses, IgM, and IgD) comparing subclinical, hospitalized, and healthy individuals during the early recovery phase of the dengue disease.***

*The values on the y-axes represent the abundance of the protein in plasma in the form of label free quantitation (LFQ) intensity. Green dot violin represents healthy donors (left); yellow dot violin represents subclinical cases (center); purple dot violin represents hospitalized patients (right). The p-values of statistical significance are shown above the respective violins.*

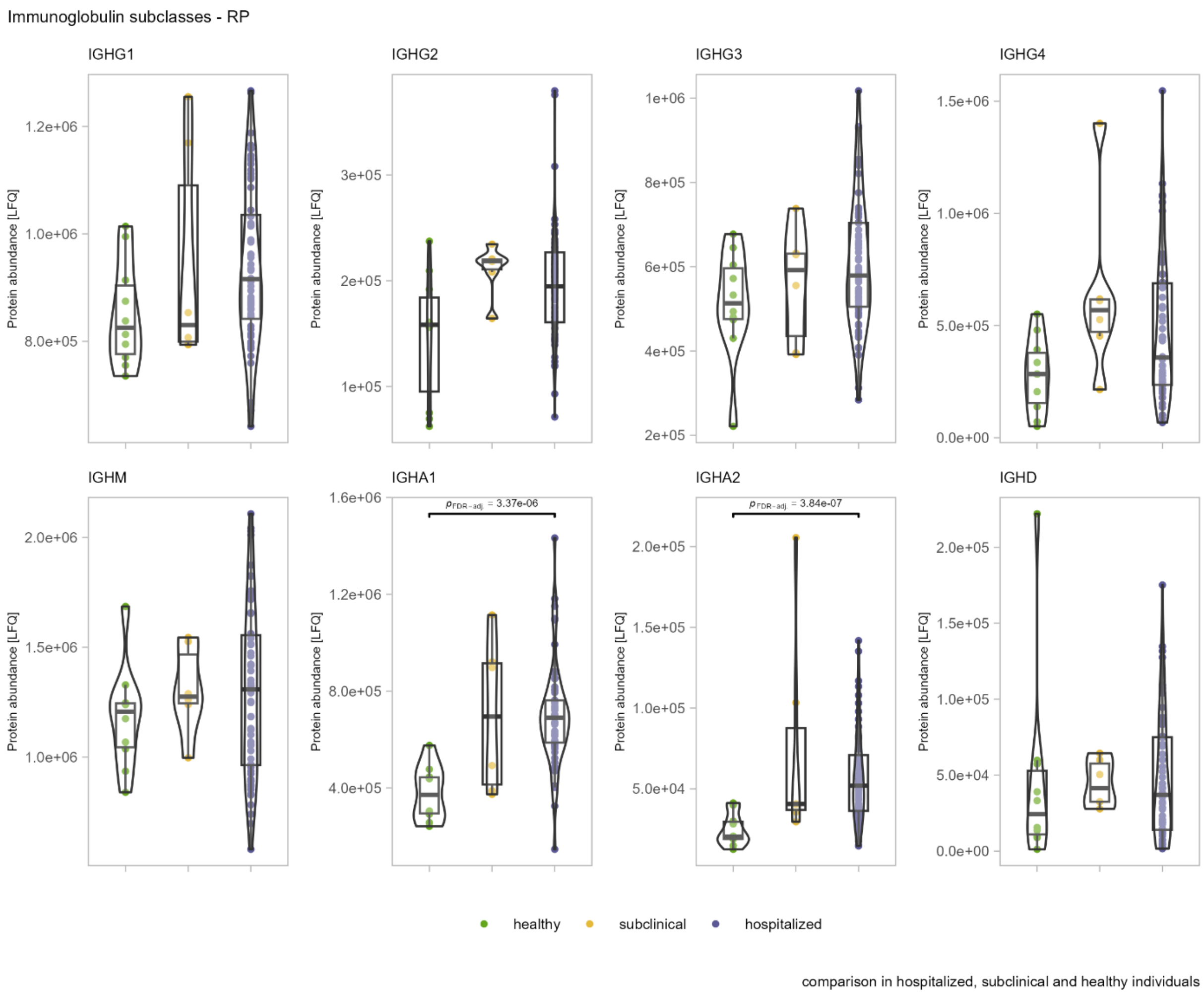


***Supplementary Figure S7. Violin plots of immunoglobulins (IgG subclasses, IgA subclasses, IgM, and IgD) comparing subclinical, hospitalized, and healthy individuals during the recovery phase of the dengue disease.***

*The values on the y-axes represent the abundance of the protein in plasma in the form of label free quantitation (LFQ) intensity. Green dot violin represents healthy donors (left); yellow dot violin represents subclinical cases (center); purple dot violin represents hospitalized patients (right). The p-values of statistical significance are shown above the respective violins.*
